# Coordinated Cytokine Responses Are Associated With Rapid HIV Reservoir Decay During Acute Treated HIV Infection

**DOI:** 10.64898/2026.08.18.26360728

**Authors:** Alton Barbehenn, Erik Lundgren, Sonia Savur, Lei Shi, Junzhe Shao, Rebecca Hoh, Heather M. Hartig, Vivian Pae, Sannidhi Sarvadhavabhatla, Sophia Donaire, Caroline H. Sheikhzadeh, Jeffrey Milush, Gregory M. Laird, Mignot Mathias, Kristen Ritter, Jeffrey Martin, Frederick Hecht, Christopher Pilcher, Stephanie E. Cohen, Susan Buchbinder, Diane Havlir, Monica Gandhi, Timothy J. Henrich, Hiroyu Hatano, Susan P. Ribeiro, Jeffrey A. Tomalka, Steven G. Deeks, Rafick P. Sekaly, Jingshen Wang, Aaron Hudson, Sulggi A. Lee

## Abstract

**Background:** The HIV reservoir is established within days of infection and persists despite antiretroviral therapy (ART). Host immune responses associated with early reservoir decay remain poorly defined.

**Methods:** We analyzed > 500 longitudinal blood samples from 67 individuals treated during acute HIV infection. Plasma cytokines and HIV reservoir size (intact and defective DNA) were quantified longitudinally. Cytokine–reservoir relationships were assessed using unsupervised clustering and complementary mixed-effects nonlinear and linear spline models; counterfactual modeling examined how cytokine concentrations at defined timepoints related to subsequent reservoir trajectories.

**Results:** TNF-α, IL-17A, and IL-10 were significantly associated with intact HIV DNA during the first 24 weeks of ART after false-discovery-rate correction. TNF-α showed one of the strongest associations with intact reservoir decay, whereas IL-10 showed the most consistent association across intact and defective HIV DNA. A coordinated cytokine cluster comprising IFN-γ, IL-10, IL-18, and TNF-α was associated with both reservoir measures. Phase-specific and counterfactual analyses further suggested that associations between several cytokines and reservoir dynamics varied with timing during early ART. These relationships persisted after adjustment for ART initiation timing, baseline viremia, initial CD4+ T-cell count, and longitudinal CD4:CD8 ratio.

**Discussion:** Coordinated inflammatory, antiviral, and immunoregulatory cytokine responses are associated with HIV reservoir decay following ART initiation during acute infection, with some relationships varying over time. These findings may inform future stage-specific HIV cure strategies that leverage the timing and balance of antiviral, inflammatory, and immunoregulatory pathways to limit reservoir persistence.

**Main Summary:** In people initiating ART during acute HIV infection, coordinated inflammatory, antiviral, and immunoregulatory cytokine responses were associated with faster HIV reservoir decay, with TNF-α and IL-10 showing particularly strong and consistent associations.

## INTRODUCTION

The HIV reservoir is established within days of infection [1–3] and persists despite suppressive antiretroviral therapy (ART), providing a source of viral rebound when treatment is interrupted. Although several studies have modeled reservoir decay during prolonged ART [4–7], only one prior study has characterized reservoir decay rates in detail during acute treated HIV [8]. Studying individuals treated during acute infection, before substantial immune exhaustion, provides an opportunity to identify host responses associated with establishment and early decay of the long-lived HIV reservoir.

The cytokine response during untreated acute HIV infection is well described [9–12], but few studies have examined cytokine changes following ART initiation [13, 14] and none in relation to HIV reservoir decay. Acute HIV induces rapid inflammatory, antiviral, and immunoregulatory responses, including IFNs, TNFs, inflammasome-associated cytokines, chemokines, and IL-10 [9–12]. One prior study related soluble markers to HIV reservoir size and identified several that remained elevated after one year of ART [10], but did not longitudinally model cytokine associations with reservoir decay or account for multiple testing [14].

Reservoir decay following ART initiation differs substantially between acute and chronic infection. We recently demonstrated ∼5-fold faster reservoir decay after ART initiation during acute infection [8] than previously described during chronic infection [15]. Here, we identified soluble immune responses associated with this accelerated reservoir decay and examined whether their timing was associated with subsequent reservoir trajectories, providing a framework for stage-specific HIV cure strategies.

## METHODS

### Study participants

Individuals with newly diagnosed acute HIV infection were enrolled in the UCSF Treat Acute HIV cohort from December 2015 to November 2020, as previously described (**Supplementary Table 1**) [8]. Participants completed monthly visits for the first 24 weeks, then every 3-4 months. The estimated date of detectable infection (EDDI) was calculated using the Infection Dating Tool [16]. All participants provided written informed consent; the UCSF institutional review board approved the study.

### HIV reservoir quantification

HIV intact and defective DNA frequencies were quantified at weeks 0, 2, 4, 8, 12, 16, 20, 24, and 52 using the intact proviral DNA assay (IPDA), as previously described [8, 17]. Genomic DNA was extracted from negatively selected CD4+ T cells (median recovery: 2×10^6^ cells; median viability: 97%) using the QIAamp DNA Mini Kit (Qiagen); samples were excluded if cell viability fell below 70%. Across >500 IPDA measurements, a median of 4.8x10^5^ CD4+ T cell genomes were interrogated per assay, with a median DNA shearing index of 0.40, and samples were batch processed alongside positive and negative controls to ensure reproducibility.

### Plasma cytokine quantification

Plasma concentrations of 23 cytokines were quantified longitudinally at the same timepoints as HIV reservoir DNA using a custom U-Plex multiplexed assay (Meso Scale Diagnostics). Fifteen cytokines meeting the prespecified quantifiability criterion were included in concentration-based analyses; assay quantifiability, handling of BLQ and unmeasured observations, and a binary quantifiability sensitivity analysis of excluded cytokines are detailed in the **Methods Supplement** and **Supplementary Tables 2-3**. Cytokine concentrations were standardized within participants and analyzed longitudinally as detailed in the **Methods Supplement**.

### Cytokine-reservoir change association modeling

Primary longitudinal analyses focused on the first 24 weeks of ART, when sampling was most frequent, with analyses through 52 weeks performed to evaluate the consistency of findings over extended follow-up. The longitudinal relationship between cytokine concentrations and reservoir size was evaluated using two complementary mixed-effects approaches with distinct purposes. **Model 1** used a biphasic linear spline with the previously identified week-5 inflection point to translate these relationships into clinically interpretable, phase-specific estimates, including percent changes in decay rate and half-lives. **Model 2**, a flexible nonlinear tensor-spline model, was used to characterize cytokine-reservoir relationships without requiring a prespecified biphasic linear form. Thus, **Model 2** provides flexibility for identifying the shape and overall evidence of association, whereas **Model 1** provides phase-based interpretation that is only directly available from the linear spline formulation (see **Methods Supplement**).

Models adjusted for pre-ART viral load, baseline CD4+ T-cell count, timing of ART initiation, and longitudinal CD4:CD8 ratio, with participant-specific random intercepts. Covariate selection and sensitivity analyses, including adjustment for baseline HIV DNA and evaluation of transformation assumptions, are described in the Methods Supplement (**Supplementary Figure 1**; **Supplementary Tables 4-5**).

Statistical significance of cytokine-reservoir associations was evaluated using nested-model deviance tests. Phase-specific effect sizes from **Model 1** were expressed as changes in decay rate and half-life per two-fold increase in cytokine concentration, as detailed in the **Methods Supplement**.

### Counterfactual modeling of subsequent reservoir trajectories

**Model 3** examined how cytokine concentrations at specified timepoints related to subsequent reservoir trajectories (**Supplementary Figure 2**). Cytokine terms were restricted to subsequent decay phases, imposing temporal ordering. The model included cytokine-by-ART-timing interaction terms, and model-based contrasts estimated differences in subsequent reservoir size associated with specified two-fold cytokine differences [18, 19]. Because the data are observational, these contrasts were not interpreted as causal effects. Full specifications and validation analyses are provided in the **Methods Supplement** [18–20].

## RESULTS

### Characteristics of study participants

A total of 67 adults with a new diagnosis of acute HIV (within ∼100 days between infection to ART initiation) were included (**Supplementary Table 1**) [8]. Median follow-up was 9.7 months, and participants achieved viral suppression by ∼30 days on dolutegravir-based therapy. Baseline HIV intact and defective DNA sizes were consistent with prior acute HIV reports [14, 21–23]. Approximately two-thirds were non-white race/ethnicity, consistent with national trends for acute HIV diagnoses [24], and prior PrEP use was common [8], reflecting San Francisco’s early adoption of PrEP [25]; sensitivity analyses excluding participants reporting PrEP use were performed.

### Plasma cytokine profiling identified baseline clinical correlations and longitudinal immune-response patterns

We profiled 23 plasma cytokines, of which 15 met the prespecified criterion for concentration-based longitudinal analyses (**Supplementary Table 2**). At ART initiation, these cytokines demonstrated distinct relationships with clinical and virologic features of acute HIV infection. Consistent with the well-described cytokine storm during acute HIV [9, 11–13], higher levels of several pro-inflammatory cytokines, including IFN-γ and IL-6, were associated with higher pre-ART plasma HIV RNA, lower initial CD4+ T cell counts, and later ART initiation relative to estimated infection (**Figure 1**). Higher levels of several pro-inflammatory cytokines were also associated with larger intact and defective HIV DNA reservoirs at ART initiation. In contrast, higher levels of immunoregulatory cytokines, including TGF-β1 and TGF-β2, were associated with smaller baseline HIV DNA reservoirs.

**Figure 1.**
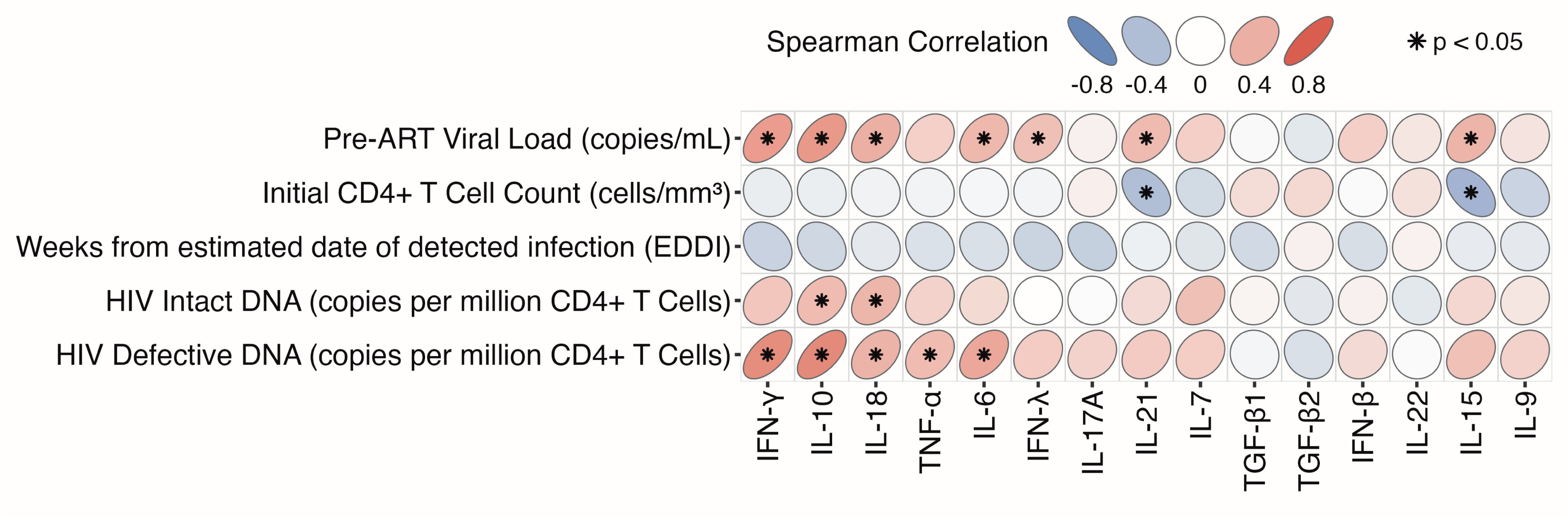
Pre-ART plasma cytokine level correlations. Spearman correlations of plasma cytokine levels at baseline (prior to ART initiation) with known clinical predictors of HIV reservoir size as well as quantified levels of HIV intact and defective DNA at baseline. Cytokines are ordered by hierarchical clustering of their correlations (see Figure 2). Stars denote p<0.05; n = 67. **Alt text:** Heatmap depicting correlations between baseline plasma cytokine levels with HIV clinical factors and HIV DNA measurements at baseline, illustrated with significance markers and ellipses with a red-blue color gradient to denote direction and magnitude of correlation.

### Longitudinal cytokine trajectories reveal coordinated inflammatory, antiviral, and immunoregulatory responses

Using repeated measures correlation, we then analyzed individual cytokine trajectories (**Supplementary Table 6**) [26]. In general, “pro-inflammatory” cytokines (e.g., IL-18) declined after ART, while more “immunoregulatory” cytokines increased (e.g., TGF-β1 and TGF-β2) (FDR q<0.05) (**Supplementary Figure 3**) [27]. Furthermore, most of the cytokines exhibited linear patterns (increasing or decreasing), although several (IL-7, IL-22, and IFN-λ) demonstrated nonlinear trajectories that varied over the first 24 weeks of ART (**Supplementary Figure 4**; **Supplementary Table 6**). Unsupervised clustering of Spearman correlations between cytokine concentration trajectories over weeks 0-24 of ART identified four distinct immune response profiles that broadly correspond to: Cluster 1 – immune cell survival, antiviral defense, and mucosal protection (IFN-β, IL-22, IL-15, IL-9); Cluster 2 – balanced antiviral responses, tissue inflammation regulation, and innate immune activation (IFN-γ, IL-10, IL-18, TNF-α); Cluster 3 – coordinated inflammation and immune regulation (IL-6, IFN-λ, IL-17A, IL-21); and Cluster 4 – immune homeostasis and tissue repair (IL-7, TGF-β1, TGF-β2) (**Figure 2**). These clusters remained consistent at individual study timepoints (**Supplementary Figure 5**).

**Figure 2.**
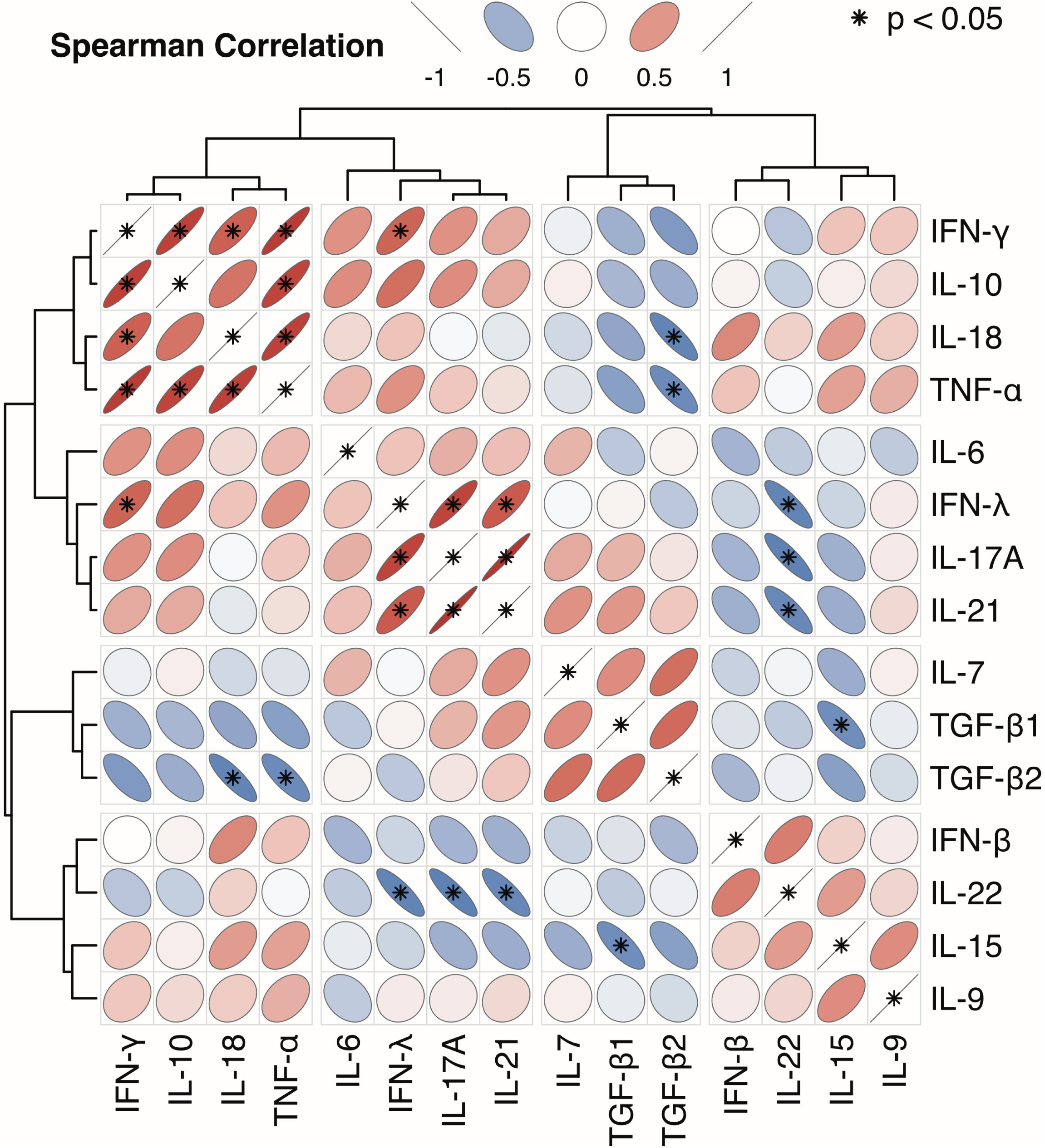
Unsupervised clustering of longitudinal plasma cytokine levels during acute treated HIV. The heatmap shows Spearman correlations between cytokine concentration trajectories over weeks 0-24 of ART. Cytokines are organized using Ward’s minimum-variance hierarchical clustering of these correlations, with the corresponding dendrogram shown above. The four resulting cytokine clusters broadly reflect distinct immune-response patterns: Cluster 1 -immune cell survival, antiviral defense, and mucosal protection (IFN-β, IL-22, IL-15, IL-9); Cluster 2 -balanced antiviral responses, tissue inflammation regulation, and innate immune activation (IFN-γ, IL-10, IL-18, TNF-α); Cluster 3 - coordinated inflammation and immune regulation (IL-6, IFN-λ, IL-17A, IL-21); and Cluster 4 - immune homeostasis and tissue repair (IL-7, TGF-β1, TGF-β2). Stars denote p<0.05. Correlations were calculated across seven longitudinal study timepoints after normalization by timepoint. **Alt text:** Heatmap showing relationships and clustering between longitudinal plasma cytokine levels, illustrated with significance markers and ellipses with a red-blue color gradient to denote direction and magnitude of correlation.

### TNF-α, IL-10, and coordinated inflammatory/antiviral responses are associated with faster intact HIV reservoir decay

Guided by observed trajectories, **Model 2** flexibly evaluated overall cytokine-reservoir relationships, while **Model 1** quantified phase-specific changes in decay and half-life using the previously identified week-5 inflection point. Using the nonlinear model, we identified several cytokines associated with intact HIV DNA during the first 24 weeks of ART. Higher IL-10, TNF-α, and IL-17A were significantly associated with longitudinal intact HIV DNA levels after correction for multiple comparisons (**Model 2**, all FDR-adjusted q<0.024; **Table 1A**). IL-18 and IFN-γ also demonstrated associations in the nonlinear model (p=0.023 and p=0.050, respectively), although these did not meet FDR-adjusted significance. These findings were generally consistent when analyses were extended through 52 weeks of ART (**Supplementary Table 7A**) and after adjustment for baseline HIV DNA levels. Moreover, the modeled associations were consistent with patterns observed in the raw longitudinal data (**Figure 3A**; **Supplementary Figure 6A**). For cytokines excluded because >30% of observations were unavailable for quantitative analysis, a separate sensitivity analysis using a binary quantifiable/BLQ indicator showed limited evidence of association with reservoir decay and was considered exploratory given the limited information captured by dichotomized measurements (**Supplementary Table 3**).

**Figure 3.**
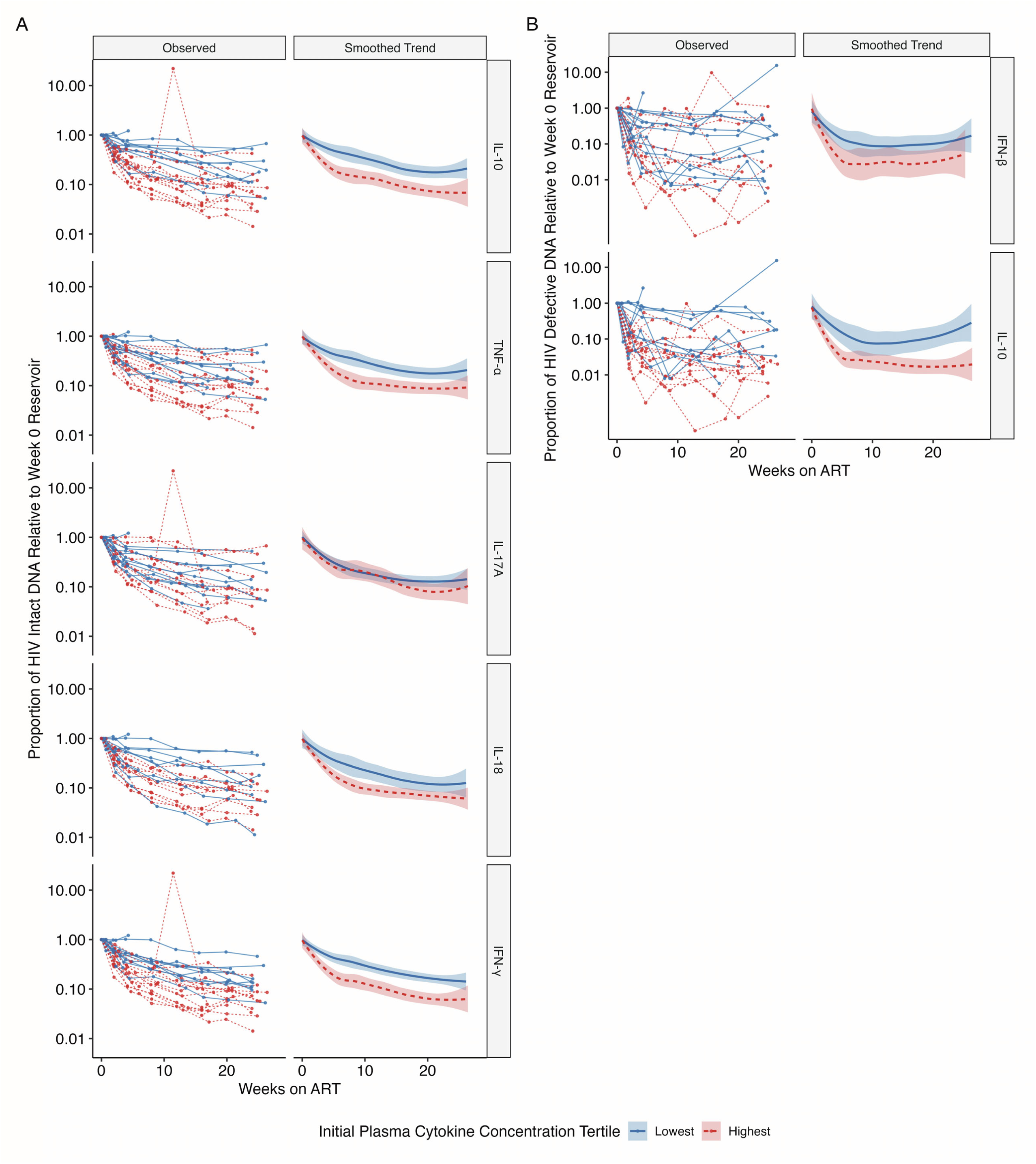
Observed HIV DNA trajectories support cytokine-reservoir associations identified in the mixed-effects models. Focusing on cytokines that showed significant or near-significant associations in the individual cytokine analyses, longitudinal HIV DNA measurements are expressed as proportional changes relative to each participant’s baseline reservoir value for HIV intact (**A**) and defective (**B**) DNA. For each cytokine, participants were grouped by their pre-ART cytokine concentration, with the highest (red dashed line) and lowest (blue solid line) tertiles shown. Left plots within each panel show participant-level reservoir trajectories; right plots show the corresponding loess-smoothed trends with shaded 95% confidence intervals. **Alt text:** Graphs comparing changes in HIV DNA measurements over time between participants with the highest and lowest plasma cytokine levels at baseline, illustrated with individual data points and smooth fitted trendlines.

**Table 1.**
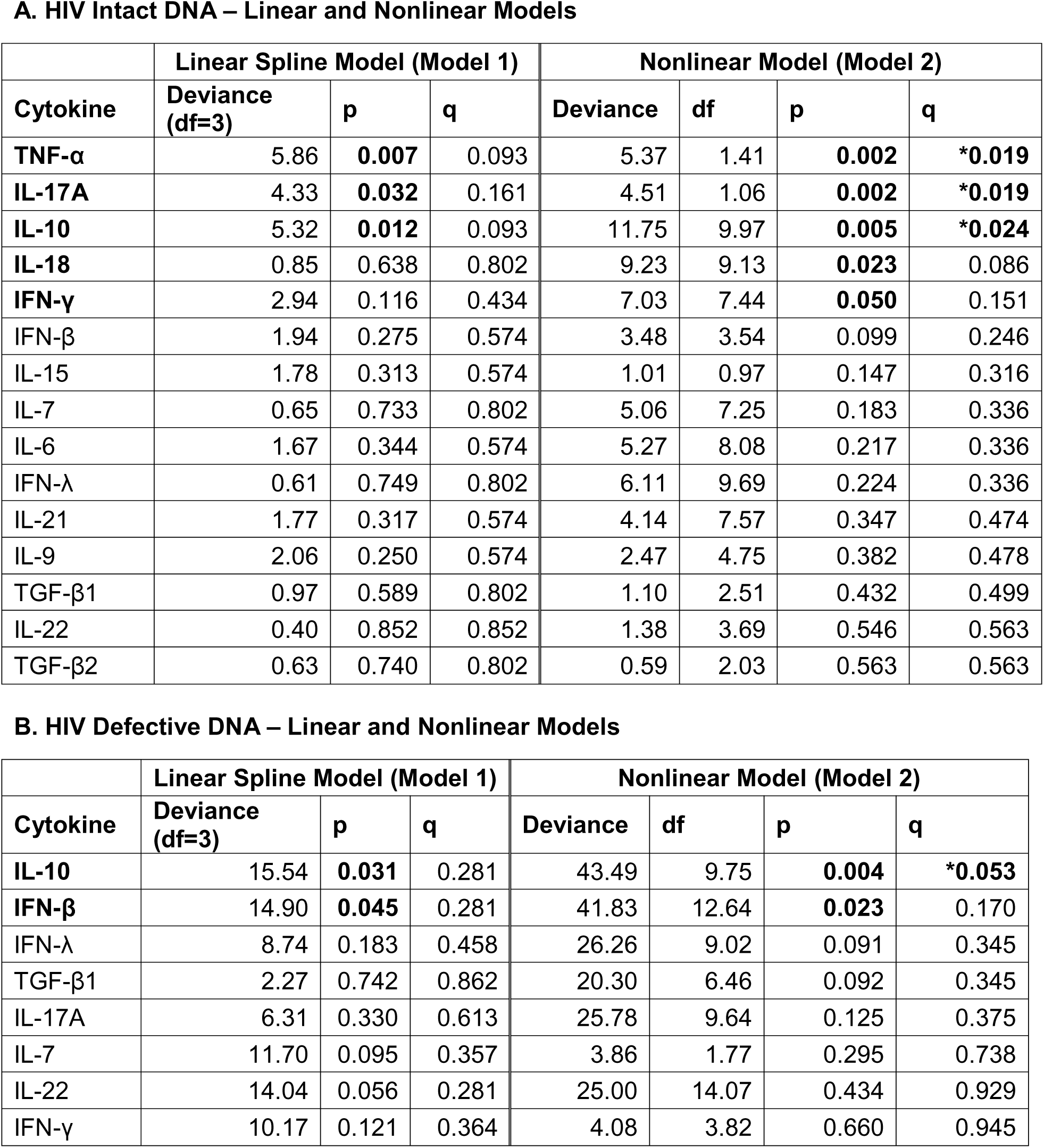
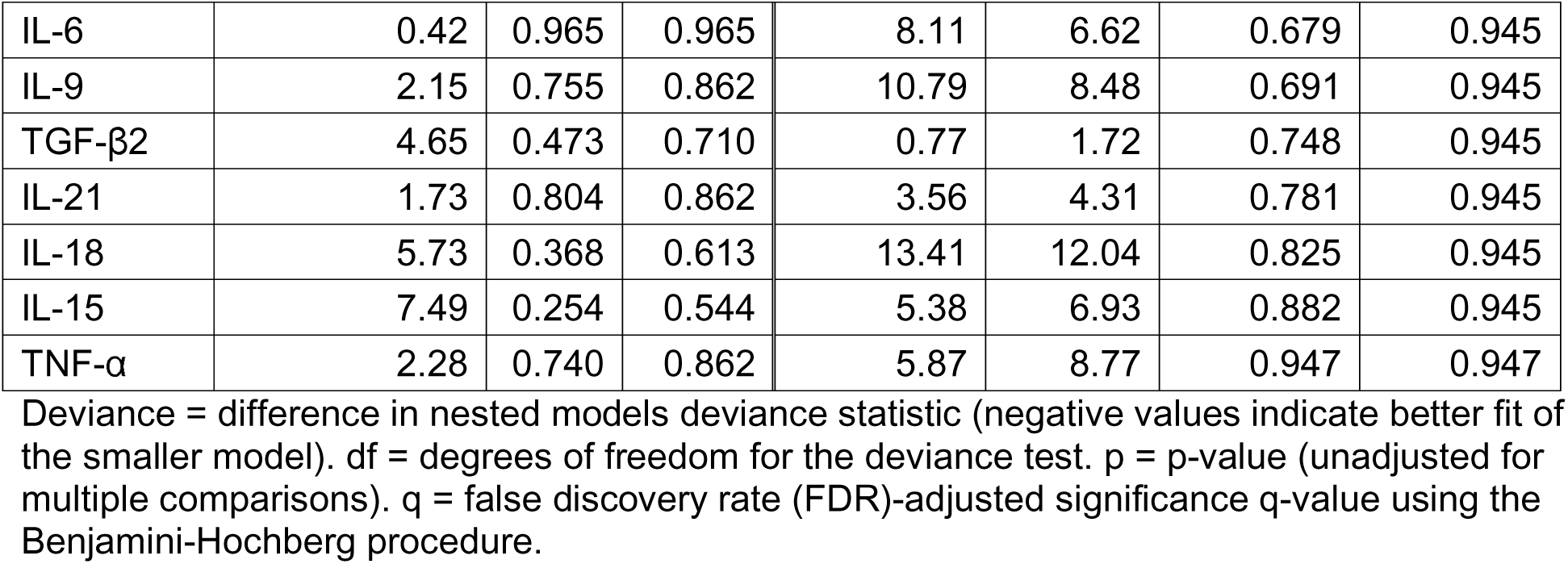
Associations between individual plasma cytokine levels and HIV reservoir size during the first 24 weeks of ART using linear and nonlinear models. We fit mixed-effect linear spline models (Model 1) and nonlinear models (Model 2) to estimate the concurrent association between each cytokine and HIV intact (**A**) and defective (**B**) DNA size during the first 24 weeks of ART. The overall association between each cytokine and HIV reservoir size is shown for both models. Rows are sorted by nonlinear model q-value. Cytokines meeting statistical significance at p<0.05 in either model are shown in bold. Statistically significant associations at FDR-adjusted q<0.05 are denoted with an asterisk.

**A. HIV Intact DNA – Linear and Nonlinear Models**
| Cytokine | Linear Spline Model (Model 1) |  |  | Nonlinear Model (Model 2) |  |  |  |
| --- | --- | --- | --- | --- | --- | --- | --- |
|  | Deviance (df=3) | p | q | Deviance | df | p | q |
| TNF-α | 5.86 | **0.007** | 0.093 | 5.37 | 1.41 | **0.002** | **\*0.019** |
| IL-17A | 4.33 | **0.032** | 0.161 | 4.51 | 1.06 | **0.002** | **\*0.019** |
| IL-10 | 5.32 | **0.012** | 0.093 | 11.75 | 9.97 | **0.005** | **\*0.024** |
| IL-18 | 0.85 | 0.638 | 0.802 | 9.23 | 9.13 | **0.023** | 0.086 |
| IFN-γ | 2.94 | 0.116 | 0.434 | 7.03 | 7.44 | **0.050** | 0.151 |
| IFN-β | 1.94 | 0.275 | 0.574 | 3.48 | 3.54 | 0.099 | 0.246 |
| IL-15 | 1.78 | 0.313 | 0.574 | 1.01 | 0.97 | 0.147 | 0.316 |
| IL-7 | 0.65 | 0.733 | 0.802 | 5.06 | 7.25 | 0.183 | 0.336 |
| IL-6 | 1.67 | 0.344 | 0.574 | 5.27 | 8.08 | 0.217 | 0.336 |
| IFN-λ | 0.61 | 0.749 | 0.802 | 6.11 | 9.69 | 0.224 | 0.336 |
| IL-21 | 1.77 | 0.317 | 0.574 | 4.14 | 7.57 | 0.347 | 0.474 |
| IL-9 | 2.06 | 0.250 | 0.574 | 2.47 | 4.75 | 0.382 | 0.478 |
| TGF-β1 | 0.97 | 0.589 | 0.802 | 1.10 | 2.51 | 0.432 | 0.499 |
| IL-22 | 0.40 | 0.852 | 0.852 | 1.38 | 3.69 | 0.546 | 0.563 |
| TGF-β2 | 0.63 | 0.740 | 0.802 | 0.59 | 2.03 | 0.563 | 0.563 |

**B. HIV Defective DNA – Linear and Nonlinear Models**
| Cytokine | Linear Spline Model (Model 1) |  |  | Nonlinear Model (Model 2) |  |  |  |
| --- | --- | --- | --- | --- | --- | --- | --- |
|  | Deviance (df=3) | p | q | Deviance | df | p | q |
| IL-10 | 15.54 | **0.031** | 0.281 | 43.49 | 9.75 | **0.004** | **\*0.053** |
| IFN-β | 14.90 | **0.045** | 0.281 | 41.83 | 12.64 | **0.023** | 0.170 |
| IFN-λ | 8.74 | 0.183 | 0.458 | 26.26 | 9.02 | 0.091 | 0.345 |
| TGF-β1 | 2.27 | 0.742 | 0.862 | 20.30 | 6.46 | 0.092 | 0.345 |
| IL-17A | 6.31 | 0.330 | 0.613 | 25.78 | 9.64 | 0.125 | 0.375 |
| IL-7 | 11.70 | 0.095 | 0.357 | 3.86 | 1.77 | 0.295 | 0.738 |
| IL-22 | 14.04 | 0.056 | 0.281 | 25.00 | 14.07 | 0.434 | 0.929 |
| IFN-γ | 10.17 | 0.121 | 0.364 | 4.08 | 3.82 | 0.660 | 0.945 |
| IL-6 | 0.42 | 0.965 | 0.965 | 8.11 | 6.62 | 0.679 | 0.945 |
| IL-9 | 2.15 | 0.755 | 0.862 | 10.79 | 8.48 | 0.691 | 0.945 |
| TGF-β2 | 4.65 | 0.473 | 0.710 | 0.77 | 1.72 | 0.748 | 0.945 |
| IL-21 | 1.73 | 0.804 | 0.862 | 3.56 | 4.31 | 0.781 | 0.945 |
| IL-18 | 5.73 | 0.368 | 0.613 | 13.41 | 12.04 | 0.825 | 0.945 |
| IL-15 | 7.49 | 0.254 | 0.544 | 5.38 | 6.93 | 0.882 | 0.945 |
| TNF-α | 2.28 | 0.740 | 0.862 | 5.87 | 8.77 | 0.947 | 0.947 |

Deviance = difference in nested models deviance statistic (negative values indicate better fit of the smaller model). df = degrees of freedom for the deviance test. p = p-value (unadjusted for multiple comparisons). q = false discovery rate (FDR)-adjusted significance q-value using the Benjamini-Hochberg procedure.

We next used the linear-spline model to determine how these cytokine associations differed across the two phases of reservoir decay. **Model 1** demonstrated that each two-fold increase in TNF-α was associated with a 23% faster decay of intact HIV DNA during the first phase (weeks 0-5) and an 11% faster decay during the second phase (weeks 5-24) (**Supplementary Table 8A**). Importantly, the phase-specific estimates also revealed that some cytokines had directionally different associations over time: higher IL-18 and IFN-γ were associated with faster intact reservoir decay during the initial phase but slower decay during the later phase (**Supplementary Table 8A**). These phase-specific findings were robust to adjustment for baseline HIV DNA levels (**Supplementary Figure 1**; **Supplementary Table 5A**). Finally, we asked whether these individual cytokine associations reflected a broader coordinated immune response. At the cluster level (**Figure 2**), cytokines in Cluster 2 (IFN-γ, IL-10, IL-18, and TNF-α) were significantly associated with HIV intact DNA levels during the first 24 weeks of ART (nonlinear model, p = 0.004) (**Table 2A**). These associations remained consistent through 52 weeks of ART despite reduced sampling density (**Supplementary Table 9A**) and were not materially affected by baseline adjustment.

**Table 2.** Associations between plasma cytokine clusters and HIV reservoir size during the first 24 weeks of ART using linear and nonlinear models. Using the four cytokine clusters identified in Figure 2, we fit mixed-effect linear spline models (Model 1) and nonlinear models (Model 2) to estimate concurrent associations between each cytokine cluster center and HIV intact (**A**) and defective (**B**) DNA size during the first 24 weeks of ART. Cluster 2 (IFN-γ, IL-10, IL-18, and TNF-α) showed the strongest and most consistent associations with longitudinal HIV intact and defective DNA. Table rows are sorted by nonlinear p-value.

**A. HIV Intact DNA – Linear and Nonlinear Model Fits**
|  | Linear Spline Model (Model 1) |  |  | Nonlinear Model (Model 2) |  |  |  |
| --- | --- | --- | --- | --- | --- | --- | --- |
| Cluster | Deviance (df=3) | p | q | Deviance | df | p | q |
| Cluster 2: IFN-γ, IL-10, IL-18, TNF-α | 3.49 | 0.070 | 0.278 | 12.48 | 10.73 | **0.004** | **\*0.016** |
| Cluster 3: IL-17A, IL-21, IFN-λ, IL-6 | 2.27 | 0.209 | 0.278 | 2.31 | 2.09 | 0.103 | 0.206 |
| Cluster 4: IL-7, TGF-β1, TGF-β2 | 2.28 | 0.207 | 0.278 | 0.70 | 1.42 | 0.345 | 0.459 |
| Cluster 1: IFN-β, IL-15, IL-22, IL-9 | 1.70 | 0.336 | 0.336 | 1.56 | 3.66 | 0.477 | 0.477 |

**B. HIV Defective DNA – Linear and Nonlinear Model Fits**
|  | Linear Spline Model (Model 1) |  |  | Nonlinear Model (Model 2) |  |  |  |
| --- | --- | --- | --- | --- | --- | --- | --- |
| Cluster | Deviance (df=3) | p | q | Deviance | df | p | q |
| Cluster 2: IFN-γ, IL-10, IL-18, TNF-α | 20.77 | **0.008** | **\*0.034** | 23.05 | 8.26 | 0.118 | 0.362 |
| Cluster 3: IL-17A, IL-21, IFN-λ, IL-6 | 11.26 | 0.108 | 0.215 | 17.57 | 6.86 | 0.181 | 0.362 |
| Cluster 4: IL-7, TGF-β1, TGF-β2 | 9.23 | 0.174 | 0.232 | 5.10 | 4.35 | 0.640 | 0.788 |
| Cluster 1: IFN-β, IL-15, IL-22, IL-9 | 2.49 | 0.717 | 0.717 | 11.30 | 9.98 | 0.788 | 0.788 |

Deviance = difference in nested models deviance statistic (negative values indicate better fit of the smaller model). df = degrees of freedom for the deviance test. p = p-value (unadjusted for multiple comparisons). q = false discovery rate (FDR)-adjusted significance q-value using the Benjamini-Hochberg procedure.

### IL-10 was the most consistent cross-reservoir predictor

Applying the same framework to HIV defective DNA, fewer significant associations were observed overall, likely reflecting greater variability in defective DNA measurements [8]. IL-10 was the only cytokine that approached significance after FDR correction (q=0.053; **Table 1B**). IFN-β was also associated with HIV defective DNA levels during first 24 weeks of ART (p=0.023), though it did not reach FDR significance and belonged to Cluster 1, which overall was not associated with HIV defective DNA (**Table 2B**; **Supplementary Table 9B**).

These findings were consistent through 52 weeks (**Supplementary Table 7B**) and with observed longitudinal patterns (**Figure 3B; Supplementary Figure 6B**). Each twofold increase in IL-10 was associated with 19% faster defective HIV DNA decay during weeks 0-5 (**Supplementary Table 8B**), with robustness to baseline adjustment (**Supplementary Figure 1; Supplementary Table 5B**). Cluster 2 (IFN-γ, IL-10, IL-18, TNF-α) was significantly associated with defective HIV DNA during the first 24 weeks (linear model, p=0.008; **Table 2B**), persisted through 52 weeks (**Supplementary Table 9B**), and was not meaningfully altered by baseline adjustment. In a multi-cytokine sensitivity analysis, relative cytokine associations remained consistent after exploratory inclusion of all 105 pairwise interactions, with IL-10 directionality most consistent with the single-cytokine findings (**Supplementary Table 10**).

### Cytokine timing was associated with subsequent reservoir trajectories

We next used counterfactual modeling (**Model 3**) to examine how cytokine concentrations at defined timepoints related to subsequent reservoir trajectories (**Supplementary Figure 2**). By restricting each cytokine measurement to reservoir outcomes occurring afterward, these models allowed us to examine whether the timing of cytokine signals was associated with subsequent reservoir decay. We modeled specified two-fold differences in cytokine concentrations at selected timepoints (e.g., weeks 0, 2, 4, or 8) and estimated subsequent reservoir trajectories. Primary analyses focused on cytokines meeting FDR-adjusted significance in the association models: IL-10, IL-17A, and TNF-α for intact HIV DNA and IL-10 for defective HIV DNA (**Table 1**; **Figure 4**). For completeness, we also examined cytokines associated at nominal significance but not after FDR correction: IL-18 and IFN-γ for intact DNA and IFN-β for defective DNA. All models adjusted for the same clinical covariates and time-varying CD4:CD8 ratio [4, 28–31].

**Figure 4.**
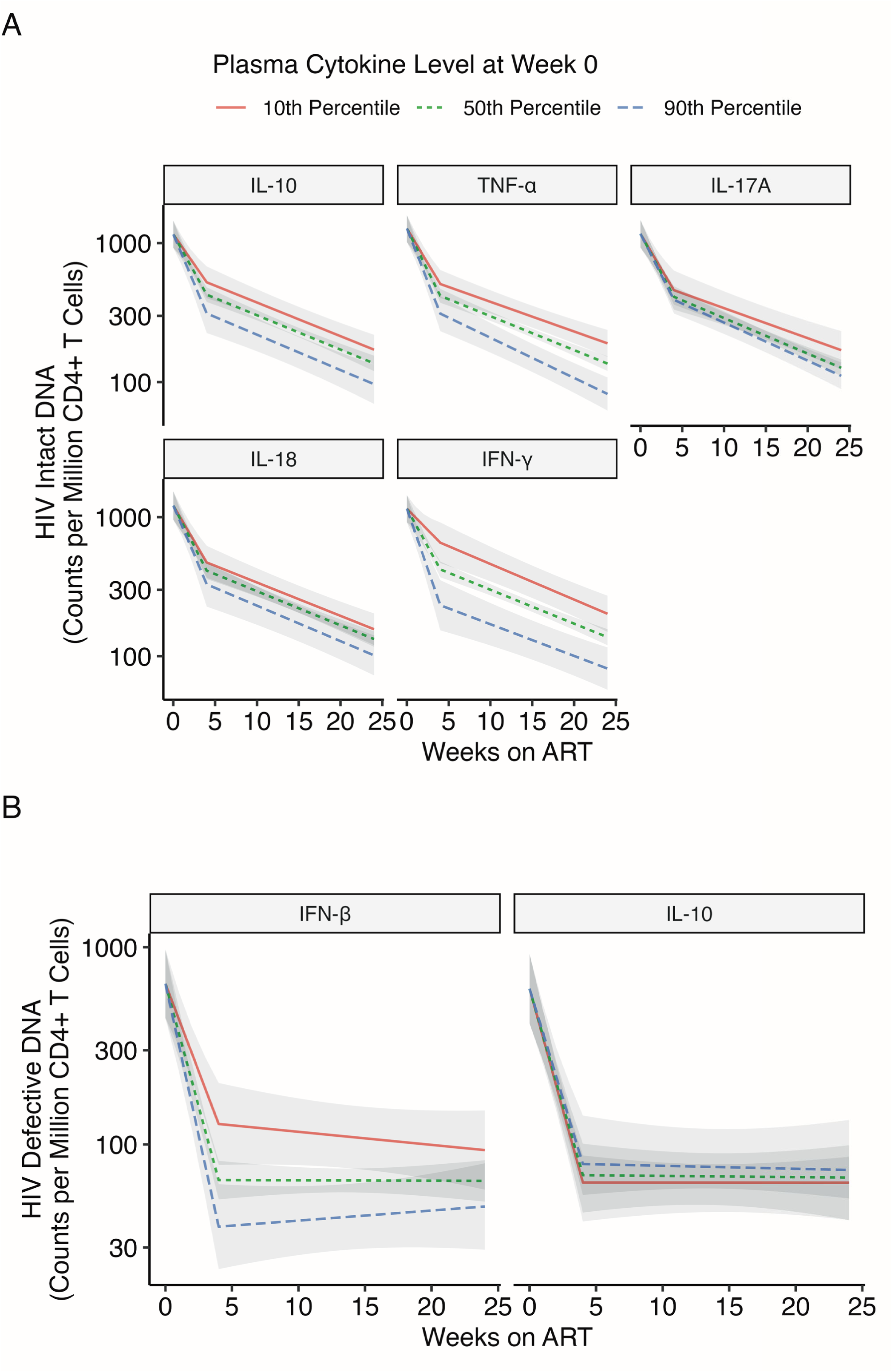
Counterfactual predicted HIV reservoir trajectories associated with specified plasma cytokine concentrations during the first 24 weeks of ART. Focusing on cytokines identified in the individual-cytokine analyses, we estimated counterfactual reservoir trajectories using Model 3. Predicted trajectories and corresponding 95% confidence intervals (grey shading) are shown for cytokine concentrations specified at the 10th, 50th, and 90th percentiles at ART initiation (week 0), for HIV intact (A) and defective (B) DNA. These estimates represent model-based counterfactual contrasts and do not by themselves establish causal effects. **Alt text:** Graphs comparing predicted HIV DNA decay trajectories over time, with lines representing different percentiles of plasma cytokine levels at baseline.

At ART initiation, model estimates comparing hypothetical cytokine concentrations at the 90^th^ versus 50^th^ or 10^th^ percentiles showed faster subsequent intact DNA decay rates for all five cytokines (**Figure 4A**). Similarly, specified two-fold higher concentrations of IL-10, IL-17A, and TNF-α at ART initiation were associated with faster initial HIV intact DNA decay during weeks 0-4 (**Supplementary Table 11A**), with generally similar patterns in analyses extending follow-up (**Supplementary Figure 7A**; **Supplementary Table 12A**). Piecewise validation models provided additional support for the direction of these temporally ordered associations (**Supplementary Table 13**). For defective DNA, estimates were less precise after week 4, consistent with the greater variability observed during the second phase (**Figures 3B** and **4B**). Higher baseline IFN-β was associated with faster subsequent defective HIV DNA decay through week 4, whereas associations at later timepoints were less consistent (**Supplementary Tables 11B-12B**; **Supplementary Figure 7B**).

We next examined how the timing of these specified cytokine differences related to subsequent reservoir size. Model-based estimates suggested that higher IL-10, TNF-α, and IL-17A concentrations were generally associated with smaller subsequent intact HIV DNA reservoirs, with the largest estimated differences observed for cytokine concentrations measured at ART initiation (**Supplementary Tables 14-15**). Notably, the direction of the estimated associations for IL-18 and IFN-γ varied by timing: higher concentrations at ART initiation predicted smaller intact HIV DNA reservoirs at 24 weeks, whereas higher concentrations at later timepoints (∼8 weeks) predicted larger reservoirs.

## DISCUSSION

In this longitudinal study of individuals initiating ART during acute HIV infection, we identified circulating cytokine profiles associated with early HIV reservoir decay. TNF-α was among the strongest individual predictors of intact HIV DNA decay, with IL-10 and IL-17A also demonstrating significant associations after FDR correction. IL-10, in contrast, was notable for the consistency of its association across both intact and defective HIV DNA outcomes. Unsupervised clustering further grouped TNF-α and IL-10 with IFN-γ and IL-18, identifying a coordinated inflammatory, antiviral, and immunoregulatory cytokine response associated with both reservoir measures. Together, these findings implicate coordinated – and potentially stage-dependent – immune responses rather than a single cytokine pathway in early reservoir dynamics.

Among these findings, TNF-α showed a particularly strong relationship with intact reservoir decay. Higher TNF-α was significantly associated with longitudinal intact HIV DNA in the flexible nonlinear model, while the phase-specific linear model estimated 23% faster intact DNA decay during weeks 0-5 and 11% faster decay during weeks 5-24 for each twofold higher TNF-α concentration. TNF-α also clustered with IFN-γ, IL-10, and IL-18 in the immune-response cluster associated with intact and defective reservoir dynamics. IL-10 showed a complementary pattern: although its association with intact HIV DNA was not stronger than that of TNF-α, it was the most consistent individual cytokine across both intact and defective HIV DNA outcomes.

IL-10’s role appears stage-specific in HIV infection. In our analysis, although IL-10 clustered with other cytokines in Cluster 2, it was the only cytokine that remained significantly associated with both intact and defective HIV DNA levels during the first 24-52 weeks of ART. These findings contrast with prior work in non-human primates, where IL-10 was linked to larger reservoirs during chronic treated SIV infection [32]. However, our unpublished data in acute treated SIV infection support a more beneficial role for IL-10 in early viral control (Rafick Sekaly, personal communication), suggesting that its effects may differ by disease stage. Mechanistically, IL-10 is classically defined as an immunoregulatory cytokine [33], known to limit inflammation, promote immune tolerance, and support regulatory T cell expansion [34–36]. However, it also exhibits context-dependent immunostimulatory properties, including enhancement of cytotoxic function and antigen-specific CD8+ T cell responses [35–41]. Human genetic studies further support this dual role: IL-10-promoting polymorphisms have been associated with higher early viral loads but improved long-term immune control [42–44]. At the same time, persistently elevated IL-10 in chronic disease settings has been linked to immune dysregulation and impaired pathogen control [45–52]. Together, these findings suggest that IL-10 exerts context-and stage-dependent effects across disease states – promoting immune control in early or controlled settings while contributing to immune dysfunction when chronically elevated (**Supplementary Figure 8**).

Other cytokines, including IFNs and TNF-α, exhibited stage-dependent effects. IFN-γ, IFN-λ, and IFN-β initially correlated with faster decay but later trends suggested slower decay during chronic phases. IL-10 and interferons both increase during acute HIV/SIV infection [11, 53–55], and regulate mucosal immunity [56, 57]. Interestingly, a prior SIV study demonstrated that while IFN treatment during chronic infection was detrimental, IFN treatment during acute infection improved viral control [54]. TNF-α, critical for pathogen control and synergizing with interferons [58, 59], can also cause tissue damage if unregulated [60, 61]. Cytokines such as IL-17, IFN-λ, and IL-18 also have dynamic roles in tissue integrity and mucosal repair [62–65], potentially aiding barrier function early while promoting damage and viral persistence when dysregulated later [64, 66, 67]. Consistent with these context-dependent effects, our models relating cytokine timing to subsequent reservoir trajectories suggested that these associations may change during the early months of ART. In particular, higher IL-18 and IFN-γ at ART initiation were associated with smaller subsequent intact HIV DNA reservoirs, whereas higher levels at later timepoints were associated with larger reservoirs (**Supplementary Figure 8**).

Our study has limitations. Although the data are observational, we extended the concurrent analyses using counterfactual models [68, 69] that related cytokine concentrations at defined timepoints to subsequent reservoir trajectories, thereby strengthening temporal ordering. These analyses provide complementary evidence beyond concurrent associations, although residual confounding precludes definitive causal inference. Although EDDI is an estimated rather than observed infection date, infection dating in this cohort benefits from the unusually comprehensive integration between UCSF and the San Francisco Department of Public Health (SFDPH), providing detailed longitudinal HIV testing histories for EDDI estimation using the validated Infection Dating Tool [16]. Nevertheless, residual dating uncertainty could influence associations during the earliest stages of infection. Plasma cytokines also capture only one layer of antiviral immunity and do not directly measure NK-cell, CD8+ T-cell, or tissue-resident effector function or identify the cellular source of soluble signals. Circulating cytokines may instead reflect a broader immune state in which early inflammatory and immunoregulatory signals influence the abundance, differentiation, trafficking, or function of cellular antiviral responses that contribute to reservoir clearance. This may also help explain the phase-dependent associations observed for several cytokines: soluble inflammatory signals may be most informative during the period of high antigen burden and rapid immune remodeling after ART initiation, while their relationship with reservoir dynamics may diminish or change as viremia is suppressed and cellular immune states evolve. Paired cellular phenotyping will therefore be important for mechanistic follow-up. Follow-up was relatively short (median 0.79 years), participants were mostly male with HIV-1 subtype B, and analyses focused on circulating cytokines rather than cell-specific markers. The HIV reservoir was quantified using IPDA, which correlates with but is less precise than full-length sequencing or viral outgrowth assays for estimating replication-competent provirus [2]. Peripheral measures largely reflect proviruses from tissue compartments [2], but tissue reservoirs were not directly assessed. Despite these limitations, the findings highlight that early, finely balanced immune responses integrating antiviral activity, regulation, and controlled inflammation are associated with rapid reservoir decay during early ART.

Acute HIV cohorts with longitudinal sampling at ART initiation are rare but uniquely positioned to define early mechanisms of viral control and reservoir establishment. By capturing immune dynamics at treatment initiation, these studies reveal pathways that shape reservoir size and persistence during a critical phase of infection. In contrast, prior HIV cure trials in chronically treated PWH have largely not achieved durable remission [70], underscoring the importance of defining determinants of reservoir formation and immune control across disease stages. Collectively, our findings provide a rationale for stage-specific cure strategies that leverage early immune signatures to inform therapeutic modulation of inflammatory and regulatory pathways in both acute and chronically treated populations, supporting durable immune-mediated HIV remission.

## Author contributions

All authors provided critical feedback in finalizing the report. S.G.D., H.H., and S.A.L. conceived and designed the study. S.G.D., H.H., and S.A.L. obtained funding to support the clinical enrollment of study participants, and S.A.L. and S.G.D. obtained funding to support characterization of the HIV reservoir. S.E.C., S.B., D.H., and M.G. facilitated coordination with San Francisco Department of Public Health and Ward 86 clinical services to link patients into care and provided critical feedback on the clinical management of acute HIV. S.A.L., R.H., S.G.D., H.M.H., V.P., S.Sar., S.D., C.H.S., and S.Sav. coordinated the collection, management, and quality control processes for the clinical data, and S.A.L., S.G.D., J.M., F.H., and C.P. provided biospecimens. J.M. and T.J.H. performed biospecimen processing. G.M.L., M.M., and K.R. performed the HIV reservoir assays. J.A.T. and R.P.S. performed the cytokine assays. G.M.L., M.M., K.R., L.S., J.S., J.W., T.J.H., A.B., and S.A.L. performed quality control analyses of the HIV reservoir assay data, and J.A.T., A.B., and S.A.L. performed quality control analyses of the cytokine assay data. A.B., S.A.L., and A.H. developed the cytokine reservoir decay models. A.B., E.L., A.H, and S.A.L. performed data visualization for the manuscript. S.A.L. and A.B. wrote the report with critical feedback from A.H. and the additional authors.

Correspondence should be addressed to S.A.L.

## Acknowledgements

The authors wish to thank all study participants of the UCSF Treat Acute HIV and SCOPE cohorts for their invaluable contributions, as well as the clinical research staff for their dedicated support throughout the study. They also gratefully acknowledge Florian Hladik, Germán G Gornalusse, and Peter Hunt for their thoughtful editorial input on the final manuscript.

## Data availability

The data underlying this article are available in the Dryad database at doi:10.5061/dryad.3tx95x6vg.

## REFERENCES

1. Whitney, J.B., et al., Rapid seeding of the viral reservoir prior to SIV viraemia in rhesus monkeys. Nature, 2014. 512(7512): p. 74-7.

2. Leyre, L., et al., Abundant HIV-infected cells in blood and tissues are rapidly cleared upon ART initiation during acute HIV infection. Sci Transl Med, 2020. 12(533).

3. Ananworanich, J., et al., Virological and immunological characteristics of HIV-infected individuals at the earliest stage of infection. J Virus Erad, 2016. 2: p. 43–48.

4. Peluso, M.J., et al., Differential decay of intact and defective proviral DNA in HIV-1-infected individuals on suppressive antiretroviral therapy. JCI Insight, 2020. 5(4).

5. Gandhi, R.T., et al., Selective Decay of Intact HIV-1 Proviral DNA on Antiretroviral Therapy. J Infect Dis, 2021. 223(2): p. 225–233.

6. Gandhi, R.T., et al., Varied Patterns of Decay of Intact Human Immunodeficiency Virus Type 1 Proviruses Over 2 Decades of Antiretroviral Therapy. J Infect Dis, 2023. 227(12): p. 1376–1380.

7. Antar, A.A., et al., Longitudinal study reveals HIV-1-infected CD4+ T cell dynamics during long-term antiretroviral therapy. J Clin Invest, 2020. 130(7): p. 3543–3559.

8. Barbehenn, A., et al., Rapid biphasic decay of intact and defective HIV DNA reservoir during acute treated HIV disease. Nat Commun, 2024. 15(1): p. 9966.

9. Stacey, A.R., et al., Induction of a striking systemic cytokine cascade prior to peak viremia in acute human immunodeficiency virus type 1 infection, in contrast to more modest and delayed responses in acute hepatitis B and C virus infections. J Virol, 2009. 83(8): p. 3719–33.

10. Hassan, A.S., et al., A Stronger Innate Immune Response During Hyperacute Human Immunodeficiency Virus Type 1 (HIV-1) Infection Is Associated With Acute Retroviral Syndrome. Clin Infect Dis, 2021. 73(5): p. 832–841.

11. Norris, P.J., et al., Elevations in IL-10, TNF-alpha, and IFN-gamma from the earliest point of HIV Type 1 infection. AIDS Res Hum Retroviruses, 2006. **22**(8): p. 757-62.

12. Muema, D.M., et al., Association between the cytokine storm, immune cell dynamics, and viral replicative capacity in hyperacute HIV infection. BMC Med, 2020. 18(1): p. 81.

13. Gilada, T., et al., Immune Activation in Primary Human Immunodeficiency Virus: Influence of Duration of Infection, Treatment, and Substance Use. Open Forum Infect Dis, 2022. 9(6): p. ofac155.

14. De Clercq, J., et al., Longitudinal patterns of inflammatory mediators after acute HIV infection correlate to intact and total reservoir. Front Immunol, 2023. 14: p. 1337316.

15. White, J.A., et al., Complex decay dynamics of HIV virions, intact and defective proviruses, and 2LTR circles following initiation of antiretroviral therapy. Proc Natl Acad Sci U S A, 2022. 119(6).

16. Grebe, E., et al., Interpreting HIV diagnostic histories into infection time estimates: analytical framework and online tool. BMC Infect Dis, 2019. 19(1): p. 894.

17. Bruner, K.M., et al., A quantitative approach for measuring the reservoir of latent HIV-1 proviruses. Nature, 2019. 566(7742): p. 120-125.

18. Lehmann, E.L. and G. Casella, *Theory of point estimation*. Second edition. ed. Springer texts in statistics. 1998, New York: Springer. 1 online resource (616 pages).

19. Arel-Bundock, V., N. Greifer, and A. Heiss, How to Interpret Statistical Models Using marginaleffects for R and Python. Journal of Statistical Software, 2024. 111(9).

20. Wood, S.N., *Generalized additive models : an introduction with R*. Second edition. ed. Chapman & Hall/CRC texts in statistical science. 2017, Boca Raton: CRC Press/Taylor & Francis Group. xx, 476 pages.

21. Ehrenberg, P.K., et al., Single-cell analyses identify monocyte gene expression profiles that influence HIV-1 reservoir size in acutely treated cohorts. Nat Commun, 2025. 16(1): p. 4975.

22. Pasternak, A.O., et al., Long-term effect of temporary ART initiated during primary HIV-1 infection on viral persistence. Nat Commun, 2025. 16(1): p. 6989.

23. Moldt, B., et al., Evaluation of HIV-1 reservoir size and broadly neutralizing antibody susceptibility in acute antiretroviral therapy-treated individuals. AIDS, 2022. 36(2): p. 205–214.

24. CDC, HIV Surveillance Supplemental Report: Estimated HIV Incidence and Prevalence in the United States, 2018–2022. https://stacks.cdc.gov/view/cdc/156513. Accessed July 13, 2025.

25. Buchbinder, S.P. and D.V. Havlir, Getting to Zero San Francisco: A Collective Impact Approach. J Acquir Immune Defic Syndr, 2019. 82 **Suppl 3**: p. S176–S182.

26. Mohr, D.L. and R.A. Marcon, Testing for a ‘within-subjects’ association in repeated measures data. Journal of Nonparametric Statistics, 2006. 17(3): p. 347–363.

27. Wood, S.N., Thin Plate Regression Splines. Journal of the Royal Statistical Society Series B: Statistical Methodology, 2003. 65(1): p. 95–114.

28. Archin, N.M., et al., Immediate antiviral therapy appears to restrict resting CD4+ cell HIV-1 infection without accelerating the decay of latent infection. Proc Natl Acad Sci U S A, 2012. 109(24): p. 9523–8.

29. Buzon, M.J., et al., Long-term antiretroviral treatment initiated at primary HIV-1 infection affects the size, composition, and decay kinetics of the reservoir of HIV-1-infected CD4 T cells. J Virol, 2014. 88(17): p. 10056–65.

30. Ananworanich, J., et al., HIV DNA Set Point is Rapidly Established in Acute HIV Infection and Dramatically Reduced by Early ART. EBioMedicine, 2016. 11: p. 68–72.

31. Crowell, T.A., et al., Virologic failure is uncommon after treatment initiation during acute HIV infection. AIDS, 2016. 30(12): p. 1943–50.

32. Harper, J., et al., Interleukin-10 contributes to reservoir establishment and persistence in SIV-infected macaques treated with antiretroviral therapy. J Clin Invest, 2022. 132(8).

33. Ouyang, W. and A. O’Garra, IL-10 Family Cytokines IL-10 and IL-22: from Basic Science to Clinical Translation. Immunity, 2019. 50(4): p. 871–891.

34. Moore, K.W., et al., Interleukin-10 and the interleukin-10 receptor. Annu Rev Immunol, 2001. 19: p. 683–765.

35. Wilke, C.M., et al., Dual biological effects of the cytokines interleukin-10 and interferon-gamma. Cancer Immunol Immunother, 2011. 60(11): p. 1529–41.

36. Groux, H., et al., Inhibitory and stimulatory effects of IL-10 on human CD8+ T cells. J Immunol, 1998. 160(7): p. 3188–93.

37. Heine, G., et al., Autocrine IL-10 promotes human B-cell differentiation into IgM- or IgG-secreting plasmablasts. Eur J Immunol, 2014. 44(6): p. 1615–21.

38. Laidlaw, B.J., et al., Interleukin-10 from CD4(+) follicular regulatory T cells promotes the germinal center response. Sci Immunol, 2017. 2(16).

39. Xi, J., et al., Stimulatory role of interleukin 10 in CD8(+) T cells through STATs in gastric cancer. Tumour Biol, 2017. 39(5): p. 1010428317706209.

40. Oft, M., Immune regulation and cytotoxic T cell activation of IL-10 agonists -Preclinical and clinical experience. Semin Immunol, 2019. 44: p. 101325.

41. Hanna, B.S., et al., Interleukin-10 receptor signaling promotes the maintenance of a PD-1(int) TCF-1(+) CD8(+) T cell population that sustains anti-tumor immunity. Immunity, 2021. 54(12): p. 2825–2841 e10.

42. Naicker, D.D., et al., Interleukin-10 promoter polymorphisms influence HIV-1 susceptibility and primary HIV-1 pathogenesis. J Infect Dis, 2009. 200(3): p. 448–52.

43. Erikstrup, C., et al., Reduced mortality and CD4 cell loss among carriers of the interleukin-10 -1082G allele in a Zimbabwean cohort of HIV-1-infected adults. AIDS, 2007. 21(17): p. 2283–91.

44. Shin, H.D., et al., Genetic restriction of HIV-1 pathogenesis to AIDS by promoter alleles of IL10. Proc Natl Acad Sci U S A, 2000. 97(26): p. 14467–72.

45. Brooks, D.G., et al., Interleukin-10 determines viral clearance or persistence in vivo. Nat Med, 2006. 12(11): p. 1301–9.

46. Ejrnaes, M., et al., Resolution of a chronic viral infection after interleukin-10 receptor blockade. J Exp Med, 2006. 203(11): p. 2461–72.

47. Roque, S., et al., IL-10 underlies distinct susceptibility of BALB/c and C57BL/6 mice to Mycobacterium avium infection and influences efficacy of antibiotic therapy. J Immunol, 2007. 178(12): p. 8028–35.

48. Omer, F.M., J.B. de Souza, and E.M. Riley, Differential induction of TGF-beta regulates proinflammatory cytokine production and determines the outcome of lethal and nonlethal Plasmodium yoelii infections. J Immunol, 2003. 171(10): p. 5430–6.

49. Belkaid, Y., et al., The role of interleukin (IL)-10 in the persistence of Leishmania major in the skin after healing and the therapeutic potential of anti-IL-10 receptor antibody for sterile cure. J Exp Med, 2001. 194(10): p. 1497–506.

50. Reed, S.G., et al., IL-10 mediates susceptibility to Trypanosoma cruzi infection. J Immunol, 1994. 153(7): p. 3135–40.

51. Couper, K.N., D.G. Blount, and E.M. Riley, IL-10: the master regulator of immunity to infection. J Immunol, 2008. 180(9): p. 5771–7.

52. Carrasco, A., et al., Mucosal Interleukin-10 depletion in steroid-refractory Crohn’s disease patients. Immun Inflamm Dis, 2022. 10(10): p. e710.

53. Gondim, M.V.P., et al., Heightened resistance to host type 1 interferons characterizes HIV-1 at transmission and after antiretroviral therapy interruption. Sci Transl Med, 2021. 13(576).

54. Sandler, N.G., et al., Type I interferon responses in rhesus macaques prevent SIV infection and slow disease progression. Nature, 2014. 511(7511): p. 601-5.

55. Teigler, J.E., et al., Distinct biomarker signatures in HIV acute infection associate with viral dynamics and reservoir size. JCI Insight, 2018. 3(10).

56. Wei, H.X., B. Wang, and B. Li, IL-10 and IL-22 in Mucosal Immunity: Driving Protection and Pathology. Front Immunol, 2020. 11: p. 1315.

57. Ye, L., D. Schnepf, and P. Staeheli, Interferon-lambda orchestrates innate and adaptive mucosal immune responses. Nat Rev Immunol, 2019. 19(10): p. 614–625.

58. Brenner, D., H. Blaser, and T.W. Mak, Regulation of tumour necrosis factor signalling: live or let die. Nat Rev Immunol, 2015. 15(6): p. 362–74.

59. Karki, R., et al., Synergism of TNF-alpha and IFN-gamma Triggers Inflammatory Cell Death, Tissue Damage, and Mortality in SARS-CoV-2 Infection and Cytokine Shock Syndromes. Cell, 2021. **184**(1): p. 149-168 e17.

60. Page, M.J., J. Bester, and E. Pretorius, The inflammatory effects of TNF-alpha and complement component 3 on coagulation. Sci Rep, 2018. 8(1): p. 1812.

61. Popa, C., et al., The role of TNF-alpha in chronic inflammatory conditions, intermediary metabolism, and cardiovascular risk. J Lipid Res, 2007. 48(4): p. 751–62.

62. Veldhoen, M., Interleukin 17 is a chief orchestrator of immunity. Nat Immunol, 2017. 18(6): p. 612–621.

63. Broggi, A., et al., IFN-lambda suppresses intestinal inflammation by non-translational regulation of neutrophil function. Nat Immunol, 2017. 18(10): p. 1084–1093.

64. Nowarski, R., et al., Epithelial IL-18 Equilibrium Controls Barrier Function in Colitis. Cell, 2015. 163(6): p. 1444–56.

65. Dupaul-Chicoine, J., et al., Control of intestinal homeostasis, colitis, and colitis-associated colorectal cancer by the inflammatory caspases. Immunity, 2010. 32(3): p. 367–78.

66. Brenchley, J.M., et al., Microbial translocation is a cause of systemic immune activation in chronic HIV infection. Nat Med, 2006. 12(12): p. 1365–71.

67. Jena, K.K., et al., Type III interferons induce pyroptosis in gut epithelial cells and impair mucosal repair. Cell, 2024. 187(26): p. 7533–7550 e23.

68. Balzer, L.B., et al., A new approach to hierarchical data analysis: Targeted maximum likelihood estimation for the causal effect of a cluster-level exposure. Stat Methods Med Res, 2019. 28(6): p. 1761–1780.

69. Nance, N., et al., The Causal Roadmap and Simulations to Improve the Rigor and Reproducibility of Real-data Applications. Epidemiology, 2024. 35(6): p. 791–800.

70. Kulpa, D.A., M. Paiardini, and G. Silvestri, Immune-mediated strategies to solving the HIV reservoir problem. Nat Rev Immunol, 2025. 25(7): p. 542–553.

