## Supplementary Tables & Figures for "Coordinated Cytokine Responses Are Associated With Rapid HIV Reservoir Decay During Acute Treated HIV Infection"

**Supplementary Table 1.** **UCSF Treat Acute HIV Study Population**. Medians (with interquartile ranges) or frequencies (with percentages) are shown.

|  | **N = 67** |
| --- | --- |
| Timing of ART initiation (days from date of estimated detected HIV infection [EDDI] to ART start date)^a^ | 29.5 (21.75 – 79.25) |
| Initial CD4+ T-cell count (cells/mm^3^) | 505 (350 – 670) |
| Pre-ART plasma HIV RNA (log_10_ copies) | 4.85 (3.69 – 5.65) |
| Age | 30.0 (25.5 – 38.0) |
| Gender (self-reported) |  |
| Male | 65 (97.0%) |
| Cisgender Female | 1 (1.50%) |
| Transgender Female | 1 (1.50%) |
| Race/ethnicity (self-reported) |  |
| White | 22 (32.8%) |
| Latinx | 20 (29.9%) |
| Asian | 14 (20.9%) |
| Black | 10 (14.9%) |
| Other | 1 (1.5%) |
| Prior pre-exposure prophylaxis (PrEP)^b^ | 29 (43.3%) |
| HIV acquired on PrEP^c^ | 6 (9.0%) |
| HIV reservoir size^d^ |  |
| HIV intact DNA | 133 (35 - 464) |
| HIV total DNA | 298 (71 - 635) |

^a^ EDDI was estimated using a validated algorithm.^15,16^ For three participants with clinical evidence of acute infection in the past 1-3 months but insufficient data for accurate EDDI (due to discordant and/or sparse testing), ART initiation was conservatively assigned to the cohort’s median number of days post-infection.

^b^ Participants with self-reported prior PrEP use (43% ever use, 20% use within 10 days of their EDDI); all modeling included sensitivity analyses with and without PrEP use participants.

^c^ Among individuals reporting PrEP use within 10 days of their EDDI, 6 participants had probable HIV acquisition while on PrEP (median baseline log_10_HIV RNA = 2.2 copies/mL) and 3 of these had M184V/I mutations.

^d^ HIV reservoir size as quantified by the intact proviral DNA assay (IPDA) after approximately 24 weeks on ART. Median copies of HIV DNA/10^6^ CD4+ T cells with interquartile ranges are shown.

**Supplementary Table 2. Quantifiability and assay availability of plasma cytokines.** Eight cytokines were excluded from the primary concentration-based analyses based on the prespecified >30% nonquantifiable/unavailable criterion (IFN-α2a, TGF-β3, IL-33, IL-8, IL-2, IL-27, IL-4, and IL-1β). For five cytokines, this primarily reflected values below the assay limit of quantification (BLQ). IL-8, IL-27, and IL-33 also had unmeasured observations because these analytes were not included in the first assay run; these observations were classified as unmeasured rather than BLQ. Percentages shown distinguish quantifiable, unmeasured, and BLQ observations within the modeling window.

| **Cytokine** | **% Quantifiable** | **% Unmeasured** | **% BLQ** |
| --- | --- | --- | --- |
| IFN-α2a | 41.9 | 0.0 | 58.1 |
| IFN-β | 83.3 | 0.0 | 16.7 |
| IFN-γ | 88.9 | 0.0 | 11.1 |
| IFN-λ | 99.8 | 0.0 | 0.2 |
| IL-10 | 86.8 | 0.0 | 13.2 |
| IL-15 | 99.8 | 0.0 | 0.2 |
| IL-17A | 76.5 | 0.0 | 23.5 |
| IL-18 | 99.8 | 0.0 | 0.2 |
| IL-1β | 67.4 | 0.0 | 32.6 |
| IL-2 | 58.4 | 0.0 | 41.6 |
| IL-21 | 96.5 | 0.0 | 3.5 |
| IL-22 | 92.8 | 0.0 | 7.2 |
| IL-27 | 63.1 | 36.9 | 0.0 |
| IL-33 | 43.3 | 36.9 | 19.8 |
| IL-4 | 65.6 | 0.0 | 34.4 |
| IL-6 | 88.7 | 0.0 | 11.3 |
| IL-7 | 98.8 | 0.0 | 1.2 |
| IL-8 | 57.1 | 36.9 | 6.0 |
| IL-9 | 88.2 | 0.0 | 11.8 |
| TGF-β1 | 95.7 | 0.0 | 4.3 |
| TGF-β2 | 91.3 | 0.0 | 8.7 |
| TGF-β3 | 42.7 | 0.0 | 57.3 |
| TNF-α | 95.1 | 0.0 | 4.9 |

**Supplementary Table 3. Binary quantifiability sensitivity analysis for cytokines excluded from concentration-based analyses.** Model 1 was refit using a binary indicator for quantifiable versus BLQ concentration for excluded cytokines with sufficient variation in quantifiability. For IL-8, IL-27, and IL-33, observations from the first assay run, in which these analytes were not measured, were treated as unmeasured and excluded rather than classified as BLQ. IL-27 could not be estimated because all observations were quantifiable when the analyte was assayed. Estimates of changes in reservoir decay slope (log2 copies per million CD4+ T cells per week; negative values indicate faster decay) with quantifiable cytokine levels, 95% confidence intervals, and p-values are shown separately for phases 1 and 2.

**A. HIV Intact DNA**

| **Cytokine** | **Phase 1 estimate** | **Phase 1 95% CI** | **Phase 1 p-value** | **Phase 2 estimate** | **Phase 2 95% CI** | **Phase 2 p-value** |
| --- | --- | --- | --- | --- | --- | --- |
| IFN-α2a | -0.20 | (-0.32, -0.07) | 0.00180 | 0.021 | (-0.008, 0.049) | 0.154 |
| IL-1β | 0.013 | (-0.116, 0.143) | 0.840 | -0.020 | (-0.050, 0.010) | 0.201 |
| IL-2 | -0.037 | (-0.160, 0.087) | 0.559 | 0.0011 | (-0.0273, 0.0295) | 0.939 |
| IL-27 | - | - | - | - | - | - |
| IL-33 | 0.068 | (-0.181, 0.317) | 0.595 | -0.042 | (-0.099, 0.015) | 0.148 |
| IL-4 | -0.081 | (-0.205, 0.044) | 0.204 | -0.00087 | (-0.03026, 0.02851) | 0.954 |
| IL-8 | 0.066 | (-0.278, 0.410) | 0.706 | -0.021 | (-0.116, 0.075) | 0.672 |
| TGF-β3 | -0.0047 | (-0.1327, 0.1233) | 0.942 | -0.0076 | (-0.0358, 0.0206) | 0.598 |

**B. HIV Defective DNA**

| **Cytokine** | **Phase 1 estimate** | **Phase 1 95% CI** | **Phase 1 p-value** | **Phase 2 estimate** | **Phase 2 95% CI** | **Phase 2 p-value** |
| --- | --- | --- | --- | --- | --- | --- |
| IFN-α2a | -0.17 | (-0.40, 0.07) | 0.159 | -0.012 | (-0.067, 0.042) | 0.654 |
| IL-1β | 0.077 | (-0.171, 0.324) | 0.544 | -0.032 | (-0.090, 0.026) | 0.277 |
| IL-2 | -0.14 | (-0.37, 0.09) | 0.242 | 0.012 | (-0.043, 0.066) | 0.675 |
| IL-27 | - | - | - | - | - | - |
| IL-33 | 0.15 | (-0.25, 0.55) | 0.457 | 0.036 | (-0.055, 0.126) | 0.440 |
| IL-4 | -0.27 | (-0.51, -0.04) | 0.0235 | 0.011 | (-0.045, 0.067) | 0.703 |
| IL-8 | -0.32 | (-0.88, 0.25) | 0.271 | 0.11 | (-0.04, 0.26) | 0.159 |
| TGF-β3 | -0.11 | (-0.35, 0.13) | 0.366 | -0.017 | (-0.070, 0.037) | 0.548 |

**Supplementary Figure 1. Robustness of cytokine effect estimates to adjustment for baseline HIV DNA levels.** Scatter plots compare estimated cytokine-associated changes in HIV DNA decay rates from models fit with and without adjustment for baseline HIV DNA. Each point represents the effect estimate for a single cytokine. The x-axis shows the percent change in decay rate per two-fold increase in cytokine concentration from models without baseline HIV DNA adjustment, while the y-axis shows corresponding estimates from models including baseline HIV DNA as both a main effect and an interaction with time on ART. The dashed diagonal line denotes y=x (perfect concordance between models). Fitted linear relationships and 95% confidence intervals are also shown. The close alignment of estimates along the diagonal indicates that adjustment for baseline HIV DNA had minimal impact on cytokine effect estimates. Greater dispersion for defective HIV DNA, particularly during phase 2 (weeks 5-24), reflects greater uncertainty in these estimates.

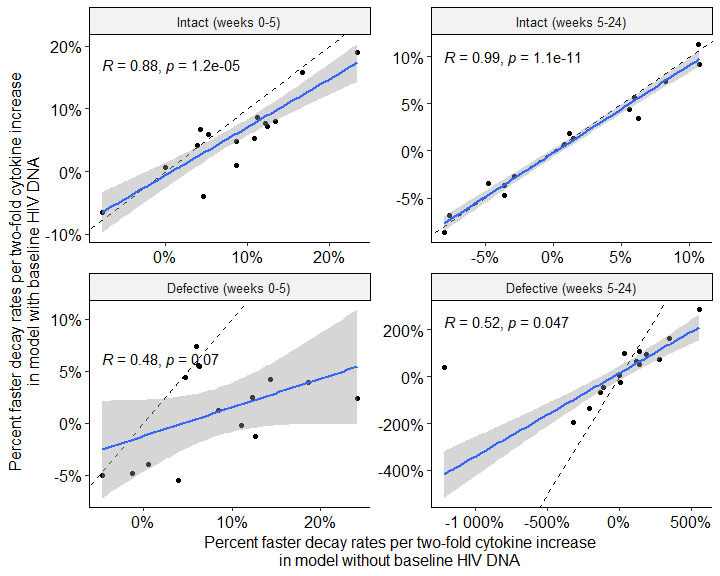

**Supplementary Table 4. CD4+ and CD8+ T cell frequencies were associated with HIV reservoir.** To test whether time-varying measures of CD4+ and/or CD8+ T cell frequencies should be in included in our final models, we tested each T cell measure (CD4+ absolute count, CD4%, CD8+ absolute count, CD8%, or CD4:CD8 ratio) in our multivariate reservoir decay models adjusted for timing of ART initiation, initial CD4+ T cell count, and pre-ART viral load. The strength of association in the longitudinal analyses was measured using a deviance test to assess the overall contribution of time-varying CD4+ and/or CD8+ T cell frequencies on HIV intact (**A**) and defective (**B**) DNA size during the first 24 weeks of ART. A different model was fit for each combination of T cell and HIV reservoir measures.

**A. HIV Intact DNA**

|  | **Overall Significance** | | **Phase 1 (Weeks 0 – 5)** | | | **Phase 2 (Weeks 5 – 24)** | | |
| --- | --- | --- | --- | --- | --- | --- | --- | --- |
| **T Cell Measure** | **Deviance (df=3)** | **p value** | **Percent Faster Decay Rate (%)** | **95% CI** | **p** | **Percent Faster Decay Rate (%)** | **95% CI** | **p** |
| CD4 Count | 10.50 | 2.43e-04 | -0.13 | (-0.24, -0.03) | 0.012 | -0.03 | (-0.11, 0.05) | 0.509 |
| CD4 Percent | 18.42 | 9.71e-08 | -4.74 | (-7.84, -1.64) | 0.003 | -2.90 | (-5.66, -0.14) | 0.040 |
| CD4:CD8 Ratio | 24.78 | 9.16e-11 | -73.64 | (-128.36, -18.93) | 0.008 | -67.17 | (-113.82, -20.52) | 0.005 |
| CD8 Count | 1.66 | 4.08e-01 | 0.01 | (-0.04, 0.05) | 0.798 | 0.04 | (-0.02, 0.11) | 0.219 |
| CD8 Percent | 12.78 | 2.80e-05 | 1.04 | (-1.27, 3.34) | 0.379 | 3.21 | (0.67, 5.75) | 0.013 |

**B. HIV Defective DNA**

|  | **Overall Significance** | | **Phase 1 (Weeks 0 – 5)** | | | **Phase 2 (Weeks 5 – 24)** | | |
| --- | --- | --- | --- | --- | --- | --- | --- | --- |
| **T Cell Measure** | **Deviance (df=3)** | **p value** | **Percent Faster Decay Rate (%)** | **95% CI** | **p** | **Percent Faster Decay Rate (%)** | **95% CI** | **p** |
| CD4 Count | 19.04 | 1.79e-02 | -0.11 | (-0.19, -0.03) | 0.009 | -0.86 | (-4.50, 2.77) | 0.641 |
| CD4 Percent | 29.82 | 1.06e-03 | -3.91 | (-6.37, -1.46) | 0.002 | -48.69 | (-204.27, 106.90) | 0.540 |
| CD4:CD8 Ratio | 27.08 | 1.88e-03 | -73.61 | (-118.10, -29.11) | 0.001 | -904.74 | (-3990.36, 2180.88) | 0.566 |
| CD8 Count | 1.72 | 8.10e-01 | 0.01 | (-0.03, 0.05) | 0.531 | 0.36 | (-0.88, 1.60) | 0.566 |
| CD8 Percent | 12.24 | 7.57e-02 | 2.01 | (0.18, 3.84) | 0.031 | 27.32 | (-44.24, 98.89) | 0.454 |

Deviance = difference in nested models deviance statistic. Percent faster decay rate is reported for each unit increase in T cell measure (i.e. a 1-count increase in CD4 count or a 1% increase in CD4 percent).

**Supplementary Table 5. Cytokine-associated changes in HIV reservoir decay rate and half-life with adjustment for baseline HIV DNA levels.** As a sensitivity analysis, Model 1 was additionally adjusted for baseline HIV DNA as a main effect and interaction with time on ART. We estimated phase-specific changes in reservoir decay rate and corresponding changes in half-life associated with each two-fold increase in plasma cytokine concentration for HIV intact (**A**) and defective (**B**) DNA during the first 24 weeks of ART. Results are shown as percent change in decay rate and change in half-life (weeks), with 95% confidence intervals and p-values. Rows are ordered to match **Table 1**.

**A. HIV Intact DNA – Fold Change Estimates from Linear Spline Model**

|  | **Phase 1 (Weeks 0-5) Change in Half-Life** | | | | **Phase 2 (Weeks 5-24) Change in Half-Life** | | | |
| --- | --- | --- | --- | --- | --- | --- | --- | --- |
| **Cytokine** | **Percent Change in Decay Rate (%)** | **Change in t_1/2_ (Weeks)** | **95% CI** | **p** | **Percent Change in Decay Rate (%)** | **Change in t_1/2_ (Weeks)** | **95% CI** | **p** |
| TNF-α | 19.0 | -0.67 | (-1.62, 0.29) | 0.170 | 11.4 | -1.58 | (-5.63, 2.48) | 0.446 |
| IL-17A | 4.8 | -0.17 | (-0.60, 0.26) | 0.447 | 5.6 | -0.80 | (-2.56, 0.97) | 0.377 |
| IL-10 | 7.2 | -0.26 | (-0.82, 0.30) | 0.360 | 7.4 | -1.02 | (-3.38, 1.34) | 0.398 |
| IL-18 | 8.0 | -0.28 | (-1.00, 0.44) | 0.449 | -8.7 | 1.23 | (-1.42, 3.87) | 0.363 |
| IFN-γ | 7.7 | -0.27 | (-0.70, 0.15) | 0.205 | -3.6 | 0.51 | (-1.17, 2.18) | 0.554 |
| IFN-β | 4.2 | -0.15 | (-0.34, 0.05) | 0.147 | -2.8 | 0.39 | (-0.45, 1.22) | 0.363 |
| IL-15 | 1.0 | -0.03 | (-1.01, 0.94) | 0.945 | 0.7 | -0.11 | (-4.39, 4.17) | 0.961 |
| IL-7 | -4.0 | 0.14 | (-0.54, 0.81) | 0.690 | 3.5 | -0.48 | (-3.14, 2.17) | 0.720 |
| IL-6 | 5.4 | -0.19 | (-0.75, 0.37) | 0.511 | 1.3 | -0.18 | (-2.46, 2.09) | 0.874 |
| IFN-λ | 6.0 | -0.21 | (-0.65, 0.23) | 0.345 | -3.4 | 0.48 | (-1.23, 2.19) | 0.581 |
| IL-21 | 8.6 | -0.30 | (-0.70, 0.10) | 0.140 | -6.9 | 0.96 | (-0.61, 2.54) | 0.231 |
| IL-9 | 15.9 | -0.55 | (-1.29, 0.19) | 0.147 | -4.7 | 0.66 | (-2.09, 3.41) | 0.639 |
| TGF-β1 | -6.5 | 0.22 | (-0.37, 0.82) | 0.459 | 9.2 | -1.30 | (-4.59, 2.00) | 0.441 |
| IL-22 | 0.7 | -0.02 | (-0.21, 0.17) | 0.814 | 1.9 | -0.26 | (-1.01, 0.50) | 0.502 |
| TGF-β2 | 6.7 | -0.23 | (-0.91, 0.45) | 0.511 | 4.4 | -0.63 | (-3.56, 2.30) | 0.673 |

**B. HIV Defective DNA – Fold Change Estimates from Linear Spline Model**

|  | **Phase 1 (Weeks 0-5) Change in Half-Life** | | | | **Phase 2 (Weeks 5-24) Change in Half-Life** | | | |
| --- | --- | --- | --- | --- | --- | --- | --- | --- |
| **Cytokine** | **Percent Change in Decay Rate (%)** | **Change in t_1/2_ (Weeks)** | **95% CI** | **p** | **Percent Change in Decay Rate (%)** | **Change in t_1/2_ (Weeks)** | **95% CI** | **p** |
| IL-10 | 3.9 | -0.07 | (-0.29, 0.15) | 0.559 | 41.0 | 55.88 | (-516, 628) | 0.848 |
| IFN-β | 5.6 | -0.09 | (-0.18, -0.01) | 0.033 | 94.6 | 112.49 | (-536, 761) | 0.734 |
| IFN-λ | -4.8 | 0.08 | (-0.08, 0.24) | 0.335 | 49.8 | 59.23 | (-358, 477) | 0.781 |
| TGF-β1 | -5.0 | 0.08 | (-0.16, 0.32) | 0.498 | -194.3 | -236.68 | (-1690, 1217) | 0.750 |
| IL-17A | 5.4 | -0.09 | (-0.27, 0.09) | 0.331 | 107.2 | 137.52 | (-764, 1039) | 0.765 |
| IL-7 | 2.5 | -0.04 | (-0.32, 0.24) | 0.774 | -137.9 | -177.96 | (-1398, 1042) | 0.775 |
| IL-22 | 4.4 | -0.07 | (-0.15, 0.01) | 0.076 | 65.8 | 58.73 | (-195, 312) | 0.650 |
| IFN-γ | -0.2 | 0.00 | (-0.16, 0.16) | 0.969 | -26.8 | -32.14 | (-331, 267) | 0.833 |
| IL-6 | -1.3 | 0.02 | (-0.20, 0.25) | 0.852 | 71.8 | 80.29 | (-437, 597) | 0.761 |
| IL-9 | -5.5 | 0.09 | (-0.20, 0.38) | 0.542 | 6.9 | 7.64 | (-308, 323) | 0.962 |
| TGF-β2 | 7.4 | -0.12 | (-0.40, 0.16) | 0.407 | -66.9 | -83.28 | (-711, 544) | 0.795 |
| IL-21 | -4.0 | 0.07 | (-0.09, 0.22) | 0.398 | -47.8 | -60.15 | (-486, 365) | 0.782 |
| IL-18 | 4.2 | -0.07 | (-0.36, 0.22) | 0.640 | 163.7 | 167.92 | (-690, 1025) | 0.701 |
| IL-15 | 2.3 | -0.04 | (-0.43, 0.36) | 0.850 | 286.1 | 312.26 | (-1382, 2007) | 0.718 |
| TNF-α | 1.3 | -0.02 | (-0.38, 0.34) | 0.911 | 97.8 | 112.01 | (-662, 886) | 0.777 |

**Supplementary Figure 2. Schematic of counterfactual Model 3 relating cytokine measurements at specified timepoints to subsequent HIV reservoir trajectories.** Model 3 uses a bi- or tri-phasic linear spline in which the cytokine concentration measured at a specified timepoint is restricted to subsequent reservoir decay phases, preventing that measurement from contributing to earlier reservoir estimates. As illustrated, a cytokine concentration measured at week 0 contributes to reservoir trajectories throughout the subsequent modeled period, whereas a concentration measured at week 4 contributes only to trajectories after week 4. For measurements at intermediate timepoints, an additional boundary is introduced to preserve this temporal ordering; for example, a week-2 cytokine measurement contributes to reservoir trajectories during weeks 2-4 and thereafter, but not during weeks 0-2. In the equations, $y_{it}$ is the reservoir size at time $t$ for participant $i$, $C_{i}$ is cytokine concentration at the specified measurement timepoint, $t_{1}=\min(t,4)$ and $t_{2}=\max(0, t-4)$ denote the illustrated decay intervals, and $\mu_{i}$ is the participant-specific random effect. These counterfactual models estimate model-based contrasts and do not by themselves establish causal effects.

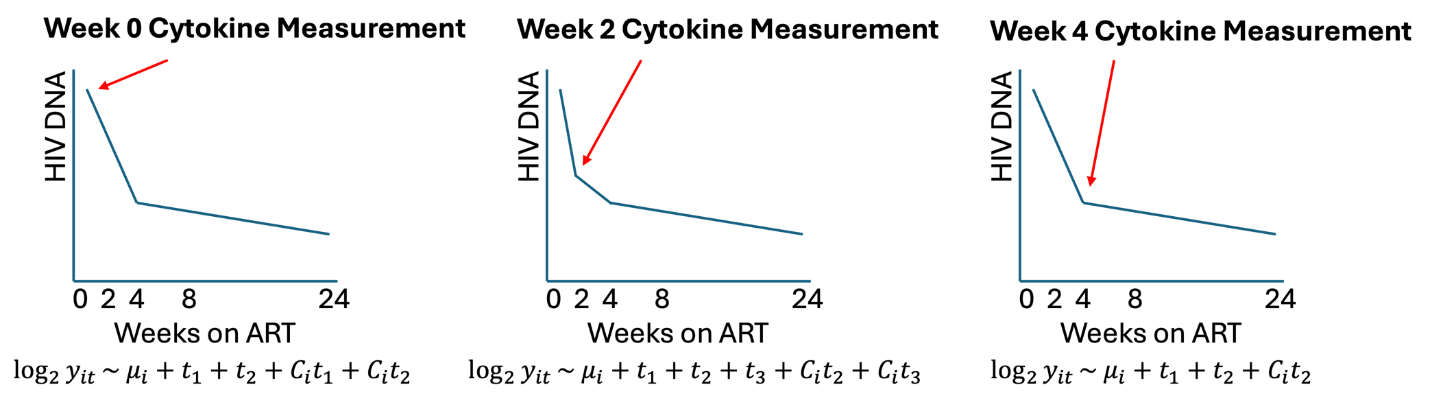

**Supplementary Table 6. Plasma cytokine concentrations in relation to time on ART.** Repeated measures Spearman correlations were calculated to demonstrate the direction and magnitude of the relationship between cytokines and time on ART (left three columns). The majority of cytokines appeared to decline over time, although many of the trends are not statistically significant (p<0.05). Mixed-effect thin plate spline models were fit to estimate nonlinearity of cytokine levels over time (fourth and fifth columns), and the degree of inter-individual variability was estimated as the proportion of total variance explained by the Gaussian random-effect term (last column). Rows are sorted by repeated measures Spearman p-value.

|  | **Repeated Measures Spearman Correlation** | | | **Mixed-Effect Thin Plate Spline** | | |
| --- | --- | --- | --- | --- | --- | --- |
| **Cytokine** | **R** | **p-value** | **q-value** | **Empirical DF** | **p-value** | **Prop. Variance Explained by Random-Effect** |
| IL-18 | -0.21 | 1.90e-05 | 0.00027 | 1 | 0.015 | 0.48 |
| TNF-α | -0.2 | 5.20e-05 | 0.00027 | 1 | 0.01 | 0.17 |
| IFN-γ | -0.2 | 5.30e-05 | 0.00027 | 1 | 0.0067 | 0.06 |
| TGF-β2 | 0.19 | 9.10e-05 | 0.00034 | 1 | 0.0026 | 0.41 |
| TGF-β1 | 0.18 | 2.70e-04 | 0.00074 | 1 | 0.0047 | 0.11 |
| IL-10 | -0.18 | 3.00e-04 | 0.00074 | 1 | 0.0094 | 0.12 |
| IL-15 | -0.11 | 2.20e-02 | 0.047 | 1 | 0.068 | 0.56 |
| IL-21 | 0.091 | 6.70e-02 | 0.13 | 1 | 0.069 | 0.21 |
| IL-7 | 0.077 | 1.20e-01 | 0.2 | 4.7 | 0.0028 | 0.35 |
| IFN-β | -0.06 | 2.30e-01 | 0.34 | 1.2 | 0.46 | 0.37 |
| IFN-λ | -0.057 | 2.50e-01 | 0.34 | 3.3 | 0.22 | 0.33 |
| IL-22 | -0.044 | 3.80e-01 | 0.47 | 4.4 | 0.078 | 0.29 |
| IL-6 | -0.039 | 4.30e-01 | 0.49 | 1 | 0.69 | 0.13 |
| IL-9 | -0.037 | 4.50e-01 | 0.49 | 1 | 0.25 | 0.18 |
| IL-17A | -0.019 | 7.00e-01 | 0.7 | 1 | 0.76 | 0.17 |

R = Repeated measures Spearman Rho. Empirical DF = empirical degrees of freedom (measures the linearity of the trend – e.g., 0 = no trend, 1 = linear trend, >1 variable trend). q-value = false discovery rate (FDR)-adjusted significance value using the Benjamini-Hochberg procedure.

**Supplementary Figure 3. Plasma cytokines demonstrate high levels of inter-individual variation.** Box-whisker plots representing observed cytokine concentrations by visit date are shown with individual trajectories in light grey lines.

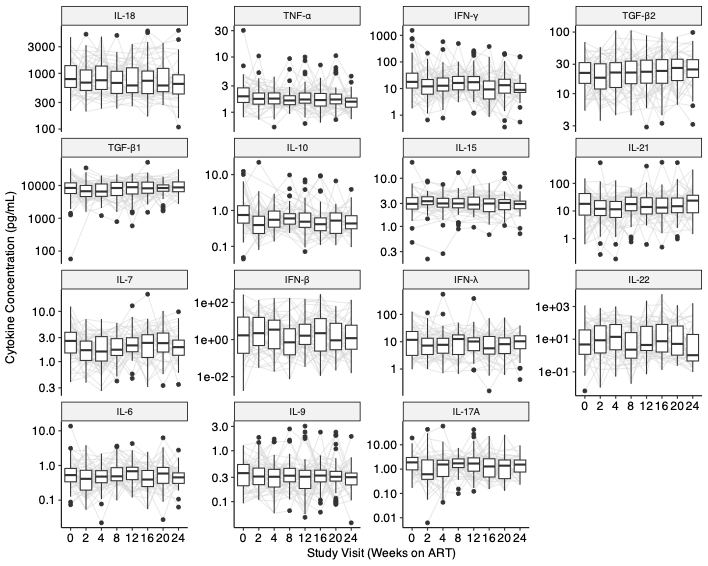

**Supplementary Figure 4. Relative trends in cytokine concentrations after ART initiation estimated using mixed-effect thin plate splines.** Raw longitudinal cytokine data for the entire cohort were used to estimate the average variability in individual cytokines over time using a method called mixed-effect thin plate splines. Mixed-effect thin plate spline trends are shown as proportional changes in cytokine levels over time (i.e., relative to cytokine level at baseline). The majority of cytokines showed linear trends (either increasing or decreasing). Three cytokines (IL-7, IL-22, and IFN-λ) were estimated to have potential nonlinear trajectories over time.

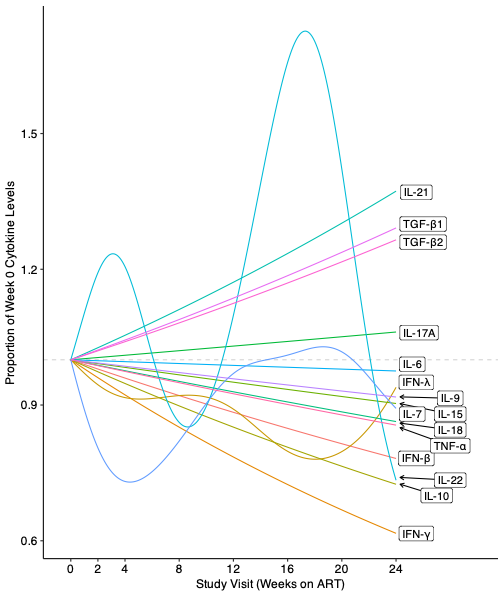

**Supplementary Figure 5. Cytokines are correlated across study visit timepoints post-ART initiation in acutely treated individuals.** Heatmaps show the Spearman correlations among plasma cytokine concentrations and markers of T cell health at each study visit. Plasma cytokines are sorted according to the hierarchical clustering used in **Figure 2**. Correlations are shown for individual cytokines (**A**) and cytokine clusters (**B**). Grey x’s denote p-values greater than 0.05; n=67.

**A.**

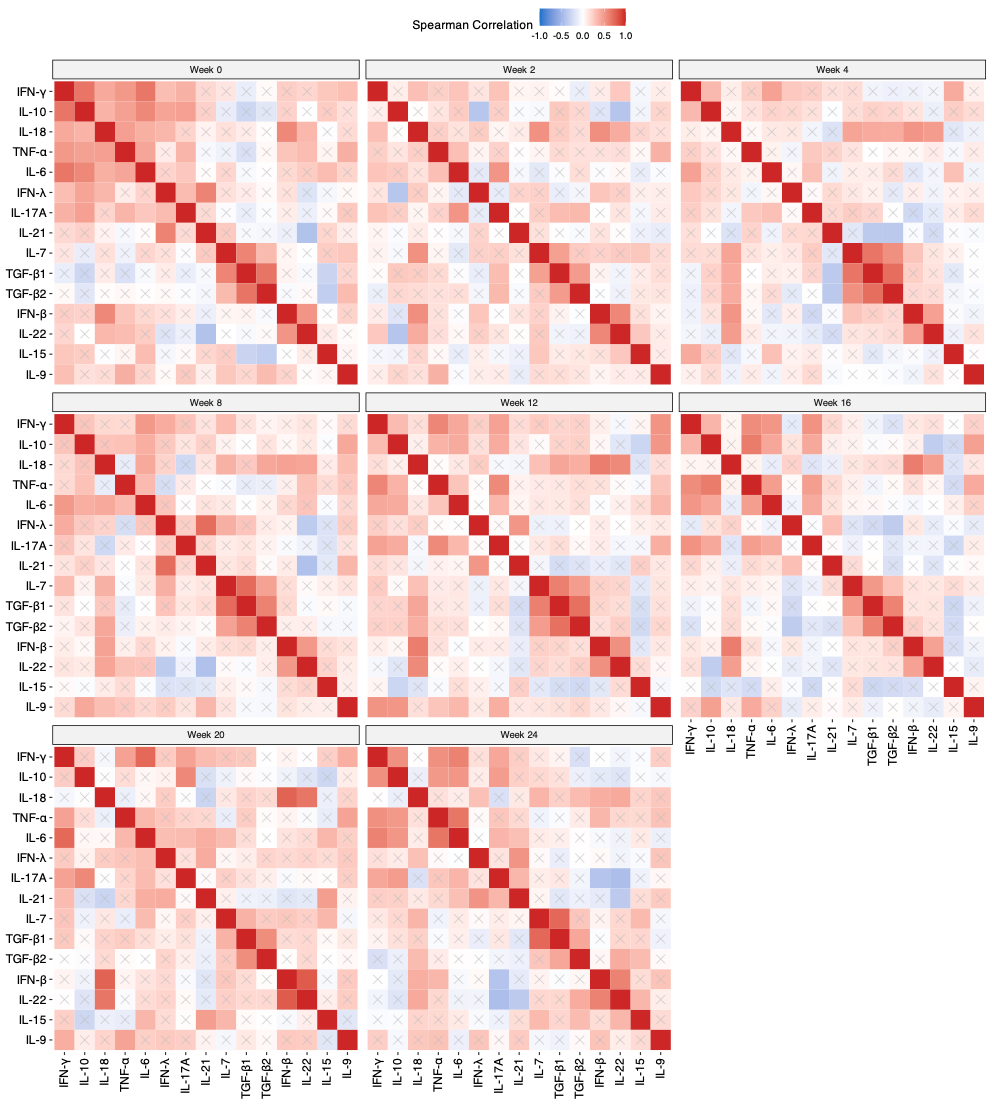

**B.**

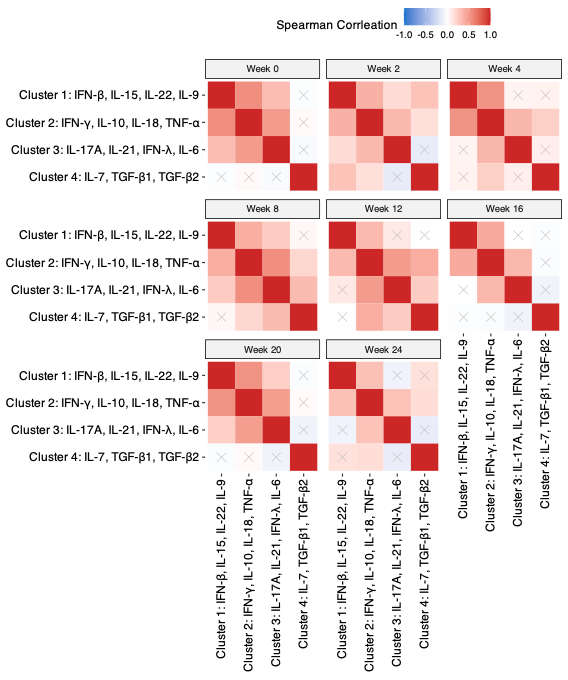

**Supplementary Table 7. Associations between individual plasma cytokine levels and HIV reservoir size during the first 52 weeks of ART using linear and nonlinear models.** We fit mixed-effect linear spline models (Model 1) and nonlinear models (Model 2) to estimate the concurrent association between each cytokine and HIV intact (**A**) and defective (**B**) DNA size during the first 52 weeks of ART. The overall association between each cytokine and HIV reservoir size is shown for both models. Rows are sorted to match **Table 1**. Cytokines meeting statistical significance at p<0.05 in either model are shown in bold. Statistically significant associations at FDR-adjusted q<0.05 are denoted with an asterisk.

**A. HIV Intact DNA – Linear and Nonlinear Models**

|  | **Linear Spline Model (Model 1)** | | | **Nonlinear Model (Model 2)** | | | |
| --- | --- | --- | --- | --- | --- | --- | --- |
| **Cytokine** | **Deviance (df=3)** | **p** | **q** | **Deviance** | **df** | **p** | **q** |
| **TNF-α** | 6.13 | **0.007** | ***0.055** | 4.99 | 2.38 | **0.010** | ***0.045** |
| **IL-17A** | 6.10 | **0.007** | ***0.055** | 5.37 | 1.34 | **0.002** | ***0.024** |
| **IL-10** | 5.60 | **0.012** | ***0.059** | 7.86 | 4.26 | **0.003** | ***0.024** |
| IL-18 | 0.65 | 0.744 | 0.797 | 7.69 | 9.83 | 0.102 | 0.209 |
| **IFN-γ** | 2.87 | 0.136 | 0.408 | 4.42 | 3.51 | **0.045** | 0.157 |
| IFN-β | 2.30 | 0.220 | 0.551 | 1.53 | -1.54 | NA | NA |
| IL-15 | 1.92 | 0.297 | 0.629 | 0.57 | 0.37 | 0.097 | 0.209 |
| IL-7 | 0.81 | 0.672 | 0.775 | 4.13 | 5.50 | 0.175 | 0.246 |
| IL-6 | 1.77 | 0.335 | 0.629 | 1.97 | 2.17 | 0.160 | 0.246 |
| IFN-λ | 0.37 | 0.872 | 0.872 | 3.60 | 3.71 | 0.104 | 0.209 |
| IL-21 | 1.15 | 0.535 | 0.775 | 0.94 | 3.16 | 0.632 | 0.632 |
| IL-9 | 3.78 | 0.062 | 0.232 | 5.03 | 6.78 | 0.165 | 0.246 |
| TGF-β1 | 0.96 | 0.610 | 0.775 | 0.69 | 2.26 | 0.569 | 0.613 |
| IL-22 | 0.86 | 0.652 | 0.775 | 1.13 | 3.12 | 0.548 | 0.613 |
| TGF-β2 | 0.82 | 0.670 | 0.775 | 1.09 | 2.15 | 0.371 | 0.473 |

**B. HIV Defective DNA – Linear and Nonlinear Models**

|  | **Linear Spline Model (Model 1)** | | | **Nonlinear Model (Model 2)** | | | |
| --- | --- | --- | --- | --- | --- | --- | --- |
| **Cytokine** | **Deviance (df=3)** | **p** | **q** | **Deviance** | **df** | **p** | **q** |
| **IL-10** | 16.33 | **0.022** | 0.337 | 54.43 | 11.41 | **0.001** | ***0.012** |
| **IFN-β** | 8.52 | 0.192 | 0.577 | 38.07 | 12.46 | **0.043** | 0.325 |
| IFN-λ | 5.79 | 0.348 | 0.746 | 16.73 | 9.45 | 0.439 | 0.822 |
| TGF-β1 | 1.99 | 0.770 | 0.831 | 25.35 | 9.94 | 0.148 | 0.410 |
| IL-17A | 6.15 | 0.321 | 0.746 | 28.19 | 10.27 | 0.105 | 0.410 |
| IL-7 | 12.08 | 0.078 | 0.403 | 7.53 | 5.91 | 0.637 | 0.875 |
| IL-22 | 4.58 | 0.469 | 0.782 | 12.61 | 4.36 | 0.159 | 0.410 |
| IFN-γ | 11.51 | 0.081 | 0.403 | 9.93 | 3.88 | 0.220 | 0.471 |
| IL-6 | 0.07 | 0.997 | 0.997 | 5.61 | 5.14 | 0.700 | 0.875 |
| IL-9 | 2.73 | 0.671 | 0.831 | 0.60 | 2.06 | 0.858 | 0.967 |
| TGF-β2 | 10.43 | 0.118 | 0.444 | 4.97 | 1.48 | 0.164 | 0.410 |
| IL-21 | 2.13 | 0.747 | 0.831 | 6.50 | 5.58 | 0.679 | 0.875 |
| IL-18 | 1.92 | 0.776 | 0.831 | 2.66 | 6.20 | 0.967 | 0.967 |
| IL-15 | 5.12 | 0.407 | 0.763 | 5.02 | 7.23 | 0.916 | 0.967 |
| TNF-α | 2.78 | 0.666 | 0.831 | 4.97 | 3.79 | 0.565 | 0.875 |

Deviance = difference in nested models deviance statistic (negative values indicate better fit of the smaller model). df = degrees of freedom for the deviance test. p = p-value (unadjusted for multiple comparisons). q = false discovery rate (FDR)-adjusted significance q-value using the Benjamini-Hochberg procedure.

**Supplementary Figure 6. Observed HIV DNA trajectories support cytokine-reservoir associations identified in the mixed-effects models.** Focusing on cytokines that showed significant or near-significant associations in the individual-cytokine analyses, raw longitudinal HIV DNA trajectories are shown for each participant as absolute values (thin grey lines). Smoothed loess trend lines for the highest and lowest tertiles of cytokine concentration are also shown (dashed and solid thick black lines, respectively), with shaded regions representing 95% confidence intervals. Observed trajectories are shown for HIV intact (**A**) and defective (**B**) DNA. These trends are generally consistent with the modeled relationships shown in Figure 3; greater variability is apparent because absolute HIV DNA levels, unlike proportional changes, retain between-participant variability in initial HIV DNA levels.

**A.**

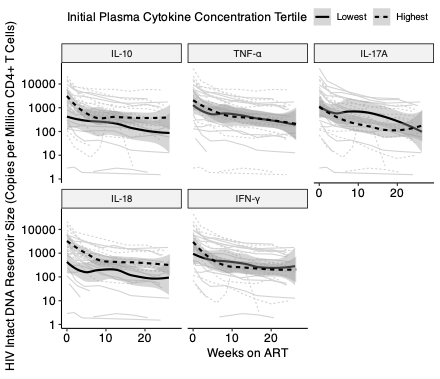

**B.**

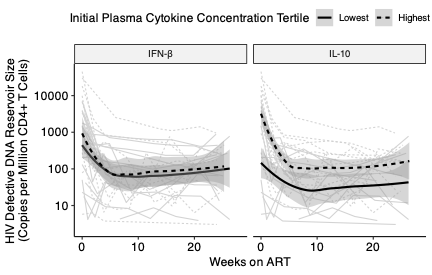

**Supplementary Table 8. Cytokine-associated changes in HIV reservoir decay rate and half-life during the first 24 weeks of ART.** Using the mixed-effect linear spline model (Model 1), we estimated phase-specific changes in reservoir decay rate and corresponding changes in half-life associated with each two-fold increase in plasma cytokine concentration for HIV intact (**A**) and defective (**B**) DNA. Results are shown as percent change in decay rate and change in half-life (weeks), with 95% confidence intervals and p-values. Rows are ordered to match **Table 1**. Cytokines meeting statistical significance at p<0.05 are shown in bold.

**A. HIV Intact DNA – Fold Change Estimates from Linear Spline Model**

|  | **Phase 1 (Weeks 0-5) Change in Half-Life** | | | | **Phase 2 (Weeks 5-24) Change in Half-Life** | | | |
| --- | --- | --- | --- | --- | --- | --- | --- | --- |
| **Cytokine** | **Percent Change in Decay Rate (%)** | **Change in t_1/2_ (Weeks)** | **95% CI** | **p** | **Percent Change in Decay Rate (%)** | **Change in t_1/2_ (Weeks)** | **95% CI** | **p** |
| TNF-α | 23.4 | -0.87 | (-2.02, 0.29) | 0.142 | 10.7 | -1.45 | (-5.50, 2.60) | 0.483 |
| IL-17A | 8.7 | -0.32 | (-0.86, 0.22) | 0.240 | 6.0 | -0.83 | (-2.62, 0.96) | 0.363 |
| IL-10 | 12.5 | -0.50 | (-1.26, 0.27) | 0.204 | 8.3 | -1.10 | (-3.45, 1.25) | 0.358 |
| IL-18 | 13.4 | -0.50 | (-1.44, 0.43) | 0.292 | -8.0 | 1.10 | (-1.53, 3.72) | 0.413 |
| IFN-γ | 12.2 | -0.48 | (-1.06, 0.10) | 0.106 | -3.6 | 0.48 | (-1.16, 2.12) | 0.563 |
| IFN-β | 3.9 | -0.14 | (-0.38, 0.09) | 0.235 | -2.8 | 0.38 | (-0.45, 1.22) | 0.367 |
| IL-15 | 8.7 | -0.33 | (-1.57, 0.91) | 0.602 | 0.9 | -0.12 | (-4.41, 4.17) | 0.956 |
| IL-7 | 4.6 | -0.17 | (-1.00, 0.66) | 0.686 | 6.3 | -0.87 | (-3.58, 1.85) | 0.532 |
| IL-6 | 10.8 | -0.41 | (-1.14, 0.32) | 0.270 | 1.5 | -0.20 | (-2.46, 2.06) | 0.862 |
| IFN-λ | 5.3 | -0.20 | (-0.72, 0.33) | 0.459 | -4.8 | 0.65 | (-1.09, 2.38) | 0.465 |
| IL-21 | 11.2 | -0.42 | (-0.95, 0.10) | 0.112 | -7.6 | 1.04 | (-0.55, 2.62) | 0.200 |
| IL-9 | 16.8 | -0.62 | (-1.52, 0.28) | 0.178 | -3.6 | 0.49 | (-2.30, 3.29) | 0.729 |
| TGF-β1 | -7.7 | 0.29 | (-0.45, 1.02) | 0.442 | 10.7 | -1.49 | (-4.86, 1.88) | 0.386 |
| IL-22 | 0.0 | 0.00 | (-0.24, 0.24) | 0.990 | 1.2 | -0.17 | (-0.92, 0.58) | 0.661 |
| TGF-β2 | 4.3 | -0.16 | (-0.99, 0.68) | 0.711 | 5.6 | -0.78 | (-3.74, 2.18) | 0.606 |

**B. HIV Defective DNA – Fold Change Estimates from Linear Spline Model**

|  | **Phase 1 (Weeks 0-5) Change in Half-Life** | | | | **Phase 2 (Weeks 5-24) Change in Half-Life** | | | |
| --- | --- | --- | --- | --- | --- | --- | --- | --- |
| **Cytokine** | **Percent Change in Decay Rate (%)** | **Change in t_1/2_ (Weeks)** | **95% CI** | **p** | **Percent Change in Decay Rate (%)** | **Change in t_1/2_ (Weeks)** | **95% CI** | **p** |
| **IL-10** | 18.6 | -0.33 | (-0.65, -0.01) | **0.043** | -1221.4 | 32176.21 | (-4.4e06, 4.5e06) | 0.989 |
| **IFN-β** | 6.2 | -0.10 | (-0.20, -0.01) | **0.034** | 186.6 | 422.85 | (-4621, 5467) | 0.869 |
| IFN-λ | -1.2 | 0.02 | (-0.16, 0.20) | 0.823 | 137.9 | 432.03 | (-7024, 7888) | 0.910 |
| TGF-β1 | -4.7 | 0.08 | (-0.18, 0.33) | 0.564 | -324.9 | -617.33 | (-6969, 5735) | 0.849 |
| IL-17A | 6.4 | -0.10 | (-0.30, 0.09) | 0.298 | 139.1 | 361.03 | (-4799, 5521) | 0.891 |
| IL-7 | 12.4 | -0.20 | (-0.51, 0.11) | 0.203 | -209.8 | -395.12 | (-4522, 3732) | 0.851 |
| IL-22 | 4.7 | -0.08 | (-0.16, 0.01) | 0.085 | 118.1 | 148.84 | (-837, 1135) | 0.767 |
| IFN-γ | 11.1 | -0.19 | (-0.40, 0.02) | 0.072 | 3.3 | 20.16 | (-6462, 6503) | 0.995 |
| IL-6 | 12.7 | -0.21 | (-0.49, 0.06) | 0.132 | 275.8 | 914.22 | (-15477, 17305) | 0.913 |
| IL-9 | 4.0 | -0.06 | (-0.38, 0.25) | 0.687 | -3.2 | -5.97 | (-1004, 992) | 0.991 |
| TGF-β2 | 6.0 | -0.10 | (-0.40, 0.21) | 0.541 | -134.3 | -277.41 | (-3498, 2943) | 0.866 |
| IL-21 | 0.5 | -0.01 | (-0.17, 0.16) | 0.920 | -114.9 | -395.43 | (-7817, 7026) | 0.917 |
| IL-18 | 14.3 | -0.24 | (-0.58, 0.11) | 0.176 | 345.2 | 611.00 | (-5155, 6377) | 0.835 |
| IL-15 | 24.2 | -0.40 | (-0.88, 0.08) | 0.102 | 551.4 | 1011.45 | (-8881, 10904) | 0.841 |
| TNF-α | 8.5 | -0.14 | (-0.54, 0.27) | 0.503 | 32.5 | 77.19 | (-2464, 2619) | 0.953 |

**Supplementary Table 9. Associations between plasma cytokine clusters and HIV reservoir size during the first 52 weeks of ART using linear and nonlinear models.** Using the four cytokine clusters identified in **Figure 2**, we fit mixed-effect linear spline models (Model 1) and nonlinear models (Model 2) to estimate concurrent associations between each cytokine cluster center and HIV intact (**A**) and defective (**B**) DNA size during the first 52 weeks of ART. Cluster 2 (IFN-γ, IL-10, IL-18, and TNF-α) showed the strongest and most consistent associations with longitudinal HIV intact and defective DNA. Table rows are sorted by nonlinear p-value. Estimates meeting statistical significance at p<0.05 in either model are shown in bold. Statistically significant associations at FDR-adjusted q<0.05 are denoted with an asterisk.

**A. HIV Intact DNA – Linear and Nonlinear Model Fits**

|  | **Linear Spline Model (Model 1)** | | | **Nonlinear Model (Model 2)** | | | |
| --- | --- | --- | --- | --- | --- | --- | --- |
| **Cluster** | **Deviance (df=3)** | **p** | **q** | **Deviance** | **df** | **p** | **q** |
| Cluster 2: IFN-γ, IL-10, IL-18, TNF-α | 3.04 | 0.118 | 0.272 | 11.26 | 10.76 | **0.013** | **0.053** |
| Cluster 3: IL-17A, IL-21, IFN-λ, IL-6 | 1.84 | 0.317 | 0.423 | 2.31 | 1.96 | 0.096 | 0.191 |
| Cluster 4: IL-7, TGF-β1, TGF-β2 | 1.45 | 0.428 | 0.428 | 0.17 | 1.47 | 0.730 | 0.730 |
| Cluster 1: IFN-β, IL-15, IL-22, IL-9 | 2.88 | 0.136 | 0.272 | 1.70 | 4.11 | 0.514 | 0.685 |

**B. HIV Defective DNA – Linear and Nonlinear Model Fits**

|  | **Linear Spline Model (Model 1)** | | | **Nonlinear Model (Model 2)** | | | |
| --- | --- | --- | --- | --- | --- | --- | --- |
| **Cluster** | **Deviance (df=3)** | **p** | **q** | **Deviance** | **df** | **p** | **q** |
| Cluster 2: IFN-γ, IL-10, IL-18, TNF-α | 19.94 | **0.009** | ***0.036** | 33.25 | 10.50 | 0.046 | 0.185 |
| Cluster 3: IL-17A, IL-21, IFN-λ, IL-6 | 12.18 | 0.079 | 0.157 | 24.46 | 10.18 | 0.184 | 0.367 |
| Cluster 4: IL-7, TGF-β1, TGF-β2 | 2.84 | 0.662 | 0.662 | 12.28 | 9.16 | 0.665 | 0.724 |
| Cluster 1: IFN-β, IL-15, IL-22, IL-9 | 4.78 | 0.449 | 0.599 | 0.81 | 1.66 | 0.724 | 0.724 |

Deviance = difference in nested models deviance statistic (negative values indicate better fit of the smaller model). df = degrees of freedom for the deviance test. p = p-value (unadjusted for multiple comparisons). q = false discovery rate (FDR)-adjusted significance q-value using the Benjamini-Hochberg procedure.

**Supplementary Table 10. Multi-cytokine ridge sensitivity analysis.** All 15 retained log2 standardized cytokines were entered simultaneously into a ridge-penalized version of Model 1. In an exploratory extension, all 105 pairwise cytokine interactions were included simultaneously. Interaction terms showed little evidence of stable pairwise effect modification. Cytokine effect rankings were highly concordant between the simultaneous 15-cytokine model and the model additionally containing all 105 pairwise interactions (Spearman correlations 0.83-0.90 across reservoir/phase combinations), indicating that the relative cytokine associations were not substantially altered by inclusion of the interaction terms. Estimates of changes in reservoir decay slope (log2 copies per million CD4+ T cells per week; negative values indicate faster decay) per standard deviation of cytokine concentration and bootstrapped 95% confidence intervals are shown.

**A. HIV Intact DNA**

|  | **Simultaneous 15-cytokine model** | | **With all pairwise interactions** | |
| --- | --- | --- | --- | --- |
| **Cytokine** | **Phase 1 estimate (95% CI)** | **Phase 2 estimate (95% CI)** | **Phase 1 estimate (95% CI)** | **Phase 2 estimate (95% CI)** |
| IFN-β | -6.6e-07 (-9.8e-05, 9.7e-05) | -1.3e-06 (-1.7e-04, 1.7e-04) | -1.7e-08 (-1.7e-05, 1.7e-05) | -3.1e-08 (-2.9e-05, 2.8e-05) |
| IFN-γ | 2.1e-08 (-3.3e-05, 3.3e-05) | 5.6e-08 (-5.7e-05, 5.7e-05) | 6.8e-09 (-1.2e-05, 1.2e-05) | 1.7e-08 (-2.0e-05, 2.0e-05) |
| IFN-λ | 4.4e-08 (-3.7e-05, 3.7e-05) | 1.2e-07 (-6.3e-05, 6.3e-05) | 5.1e-08 (-3.4e-05, 3.4e-05) | 1.3e-07 (-5.8e-05, 5.8e-05) |
| IL-10 | -0.0011 (-0.0056, 0.0033) | -0.0031 (-0.0095, 0.0032) | -0.0010 (-0.0069, 0.0048) | -0.0031 (-0.0104, 0.0042) |
| IL-15 | 6.2e-08 (-4.0e-05, 4.0e-05) | 1.3e-07 (-6.7e-05, 6.7e-05) | 0.0010 (-0.0036, 0.0057) | 0.0022 (-0.0041, 0.0084) |
| IL-17A | -3.8e-07 (-9.9e-05, 9.8e-05) | -1.2e-06 (-1.7e-04, 1.7e-04) | -9.4e-08 (-5.6e-05, 5.5e-05) | -3.2e-07 (-9.6e-05, 9.5e-05) |
| IL-18 | 2.4e-09 (-4.8e-05, 4.8e-05) | 4.4e-08 (-8.2e-05, 8.2e-05) | 7.2e-10 (-6.4e-06, 6.4e-06) | 2.6e-09 (-1.1e-05, 1.1e-05) |
| IL-21 | 1.3e-08 (-4.7e-05, 4.7e-05) | 7.5e-08 (-7.9e-05, 8.0e-05) | 2.3e-09 (-1.2e-05, 1.2e-05) | 8.2e-09 (-2.1e-05, 2.1e-05) |
| IL-22 | -3.6e-07 (-7.6e-05, 7.5e-05) | -8.5e-07 (-1.3e-04, 1.3e-04) | -4.3e-04 (-3.7e-03, 2.8e-03) | -9.9e-04 (-5.9e-03, 3.9e-03) |
| IL-6 | -6.4e-08 (-4.6e-05, 4.6e-05) | -1.7e-07 (-7.9e-05, 7.9e-05) | -1.3e-09 (-1.2e-05, 1.2e-05) | -4.2e-09 (-2.1e-05, 2.1e-05) |
| IL-7 | -6.2e-07 (-1.0e-04, 1.0e-04) | -1.6e-06 (-1.7e-04, 1.7e-04) | -2.8e-09 (-9.4e-06, 9.4e-06) | -7.1e-09 (-1.6e-05, 1.6e-05) |
| IL-9 | 3.7e-08 (-5.0e-05, 5.0e-05) | 8.7e-08 (-8.6e-05, 8.7e-05) | 2.1e-09 (-6.7e-06, 6.7e-06) | 5.1e-09 (-1.2e-05, 1.2e-05) |
| TGF-β1 | -6.9e-07 (-1.2e-04, 1.1e-04) | -1.8e-06 (-2.0e-04, 2.0e-04) | -1.1e-08 (-1.5e-05, 1.5e-05) | -2.7e-08 (-2.6e-05, 2.6e-05) |
| TGF-β2 | -1.0e-06 (-1.3e-04, 1.2e-04) | -2.6e-06 (-2.2e-04, 2.1e-04) | -1.1e-08 (-1.4e-05, 1.4e-05) | -2.6e-08 (-2.4e-05, 2.4e-05) |
| TNF-α | -2.5e-05 (-6.6e-04, 6.0e-04) | -6.7e-05 (-1.1e-03, 1.0e-03) | -2.8e-08 (-2.4e-05, 2.3e-05) | -7.3e-08 (-4.0e-05, 4.0e-05) |

**B. HIV Defective DNA**

|  | **Simultaneous 15-cytokine model** | | **With all pairwise interactions** | |
| --- | --- | --- | --- | --- |
| **Cytokine** | **Phase 1 estimate (95% CI)** | **Phase 2 estimate (95% CI)** | **Phase 1 estimate (95% CI)** | **Phase 2 estimate (95% CI)** |
| IFN-β | -4.4e-08 (-4.6e-05, 4.6e-05) | -5.9e-08 (-7.8e-05, 7.8e-05) | -4.8e-09 (-1.6e-05, 1.6e-05) | -5.2e-09 (-2.8e-05, 2.8e-05) |
| IFN-γ | -9.0e-08 (-5.9e-05, 5.9e-05) | -2.3e-07 (-1.0e-04, 1.0e-04) | -6.1e-09 (-1.8e-05, 1.8e-05) | -1.6e-08 (-3.0e-05, 3.0e-05) |
| IFN-λ | 0.0065 (-0.0068, 0.0197) | 0.013 (-0.001, 0.027) | 0.0018 (-0.0038, 0.0074) | 0.0039 (-0.0047, 0.0126) |
| IL-10 | -1.4e-07 (-7.5e-05, 7.5e-05) | -3.4e-07 (-1.3e-04, 1.3e-04) | -1.3e-08 (-2.6e-05, 2.6e-05) | -3.1e-08 (-4.5e-05, 4.5e-05) |
| IL-15 | 1.8e-08 (-4.0e-05, 4.0e-05) | 6.3e-08 (-6.7e-05, 6.7e-05) | 6.8e-09 (-2.4e-05, 2.4e-05) | 2.3e-08 (-4.1e-05, 4.1e-05) |
| IL-17A | 2.0e-07 (-8.4e-05, 8.5e-05) | 4.5e-07 (-1.4e-04, 1.5e-04) | 1.1e-07 (-5.6e-05, 5.7e-05) | 2.5e-07 (-9.7e-05, 9.7e-05) |
| IL-18 | 5.6e-08 (-5.9e-05, 5.9e-05) | 1.7e-07 (-1.0e-04, 1.0e-04) | 0.0026 (-0.0097, 0.0150) | 0.0083 (-0.0063, 0.0230) |
| IL-21 | -4.2e-08 (-6.3e-05, 6.3e-05) | -1.5e-07 (-1.1e-04, 1.1e-04) | -6.1e-09 (-4.9e-05, 4.9e-05) | -4.1e-08 (-8.3e-05, 8.3e-05) |
| IL-22 | 4.1e-07 (-1.2e-04, 1.2e-04) | 1.3e-06 (-2.0e-04, 2.0e-04) | 6.9e-08 (-4.9e-05, 4.9e-05) | 2.2e-07 (-8.3e-05, 8.4e-05) |
| IL-6 | -1.7e-08 (-4.9e-05, 4.9e-05) | -3.5e-08 (-8.4e-05, 8.4e-05) | -5.2e-09 (-3.7e-05, 3.7e-05) | -9.7e-09 (-6.3e-05, 6.3e-05) |
| IL-7 | -0.0013 (-0.0075, 0.0049) | -0.0036 (-0.0130, 0.0058) | -0.0032 (-0.0169, 0.0105) | -0.011 (-0.026, 0.004) |
| IL-9 | 3.1e-08 (-3.4e-05, 3.4e-05) | 6.3e-08 (-5.9e-05, 5.9e-05) | 1.7e-08 (-2.3e-05, 2.3e-05) | 3.4e-08 (-4.0e-05, 4.0e-05) |
| TGF-β1 | 1.7e-08 (-6.1e-05, 6.1e-05) | 3.2e-09 (-1.1e-04, 1.1e-04) | 2.3e-08 (-5.8e-05, 5.8e-05) | 1.7e-08 (-1.0e-04, 1.0e-04) |
| TGF-β2 | 8.9e-09 (-5.4e-05, 5.4e-05) | 6.3e-10 (-9.2e-05, 9.2e-05) | 2.8e-05 (-2.0e-03, 2.0e-03) | 3.4e-05 (-3.4e-03, 3.4e-03) |
| TNF-α | -1.1e-06 (-1.7e-04, 1.7e-04) | -2.6e-06 (-2.9e-04, 2.8e-04) | -6.5e-08 (-4.2e-05, 4.2e-05) | -1.5e-07 (-7.1e-05, 7.1e-05) |

**Supplementary Table 11. Counterfactual estimates of changes in HIV reservoir decay rate associated with specified two-fold differences in plasma cytokine concentrations during the first 24 weeks of ART**. Using Model 3, we estimated changes in subsequent reservoir decay rate associated with a two-fold difference in cytokine concentration at specified measurement timepoints. Analyses focused on IL-10, TNF-α, IL-17A, IL-18, and IFN-γ for intact HIV DNA (**A**) and IFN-β and IL-10 for defective HIV DNA (**B**), based on associations identified in Models 1 and 2. Values are estimated changes in reservoir decay slope (log2 copies per million CD4+ T cells per week; negative values indicate faster decay) with 95% confidence intervals and p-values. Estimates with p<0.05 are shown in bold.

**A. HIV Intact DNA – first 24 weeks on ART**

| **Cytokine Measurement Timepoint** | **Subsequent  Reservoir Interval** | **IL-10** | **TNF-α** | **IL-17A** | **IL-18** | **IFN-γ** |
| --- | --- | --- | --- | --- | --- | --- |
| Week 0 | Weeks 0-4 | -0.04 (-0.086, 0.007); 0.095 | -0.091 (-0.19, 0.005); 0.064 | -0.016 (-0.074, 0.042); 0.59 | -0.054 (-0.15, 0.04); 0.26 | **-0.071 (-0.12,  -0.021); 0.006** |
|  | Weeks 4-24 | -0.001 (-0.01, 0.009); 0.87 | -0.013 (-0.032, 0.005); 0.16 | -0.007 (-0.018, 0.004); 0.24 | -0.002 (-0.019, 0.014); 0.77 | 0.002 (-0.007, 0.011); 0.64 |
| Week 2 | Weeks 2-4 | 0.015 (-0.08, 0.11); 0.76 | -0.1 (-0.29, 0.088); 0.3 | -0.039 (-0.097, 0.019); 0.19 | -0.06 (-0.2, 0.083); 0.41 | 0.02 (-0.055, 0.095); 0.6 |
|  | Weeks 4-24 | -0.005 (-0.017, 0.006); 0.36 | 0.002 (-0.022, 0.025); 0.89 | -0.001 (-0.007, 0.006); 0.84 | 0.004 (-0.013, 0.022); 0.63 | 0.004 (-0.005, 0.012); 0.37 |
| Week 4 | Weeks 4-24 | -0.002 (-0.012, 0.007); 0.63 | -0.009 (-0.026, 0.007); 0.27 | **-0.006 (-0.011,  -0.001); 0.027** | 0.006 (-0.003, 0.016); 0.2 | 0.003 (-0.003, 0.009); 0.29 |
| Week 8 | Weeks 8-24 | -0.007 (-0.026, 0.011); 0.42 | -0.009 (-0.039, 0.021); 0.57 | 0.006 (-0.009, 0.021); 0.45 | -0.002 (-0.021, 0.017); 0.8 | 0.005 (-0.008, 0.018); 0.47 |
| Week 12 | Weeks 12-24 | -0.001 (-0.016, 0.013); 0.85 | 0.006 (-0.027, 0.038); 0.74 | 0.009 (-0.003, 0.02); 0.14 | 0.003 (-0.017, 0.024); 0.77 | 0.004 (-0.008, 0.016); 0.51 |
| Week 16 | Weeks 16-24 | -0.008 (-0.033, 0.017); 0.52 | -0.002 (-0.046, 0.041); 0.92 | -0.004 (-0.024, 0.015); 0.66 | 0.015 (-0.013, 0.043); 0.31 | 0.01 (-0.012, 0.031); 0.37 |
| Week 20 | Weeks 20-24 | -0.004 (-0.043, 0.034); 0.82 | 0.011 (-0.047, 0.069); 0.72 | -0.003 (-0.032, 0.027); 0.86 | 0.06 (-0.003, 0.12); 0.062 | 0.009 (-0.015, 0.032); 0.48 |

**B. HIV Defective DNA – first 24 weeks on ART**

| **Cytokine Measurement Timepoint** | **Subsequent**  **Reservoir Interval** | **IFN-β** | **IL-10** |
| --- | --- | --- | --- |
| Week 0 | Weeks 0-4 | **-0.038 (-0.069, -0.008); 0.014** | 0.027 (-0.052, 0.11); 0.51 |
|  | Weeks 4-24 | 0.004 (-0.003, 0.01); 0.32 | -0.002 (-0.02, 0.016); 0.84 |
| Week 2 | Weeks 2-4 | -0.014 (-0.082, 0.054); 0.69 | 0.12 (-0.034, 0.28); 0.13 |
|  | Weeks 4-24 | 0.003 (-0.006, 0.011); 0.56 | -0.008 (-0.029, 0.013); 0.47 |
| Week 4 | Weeks 4-24 | 0.004 (-0.002, 0.01); 0.15 | -0.017 (-0.038, 0.004); 0.11 |
| Week 8 | Weeks 8-24 | 0.001 (-0.008, 0.01); 0.8 | 0.007 (-0.023, 0.037); 0.66 |
| Week 12 | Weeks 12-24 | -0.003 (-0.016, 0.011); 0.7 | -0.003 (-0.028, 0.021); 0.8 |
| Week 16 | Weeks 16-24 | **0.015 (0.001, 0.03); 0.038** | -0.04 (-0.085, 0.004); 0.079 |
| Week 20 | Weeks 20-24 | 0.011 (-0.023, 0.046); 0.52 | 0.02 (-0.062, 0.1); 0.64 |

**Supplementary Figure 7. Counterfactual predicted HIV reservoir trajectories associated with specified plasma cytokine concentrations during the first 52 weeks of ART.** Focusing on cytokines identified in the individual cytokine analyses, we estimated counterfactual reservoir trajectories using Model 3. Predicted trajectories and corresponding 95% confidence intervals (grey shading) are shown for cytokine concentrations specified at the 10th (red), 50th (green), and 90th (blue) percentiles at ART initiation (week 0), for HIV intact (**A**) and defective (**B**) DNA. These estimates represent model-based counterfactual contrasts and do not by themselves establish causal effects.

**A.**

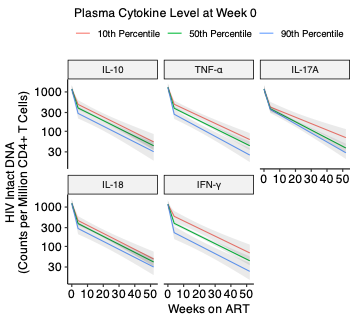

**B.**

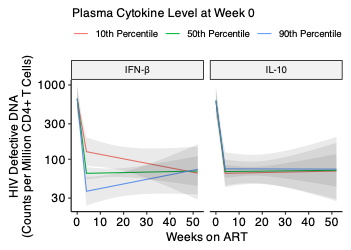

**Supplementary Table 12. Counterfactual estimates of changes in HIV reservoir decay rate associated with specified two-fold differences in plasma cytokine concentrations during the first 52 weeks of ART.** Using Model 3, we estimated changes in subsequent reservoir decay rate associated with a two-fold difference in cytokine concentration at specified measurement timepoints. Analyses focused on IL-10, TNF-α, IL-17A, IL-18, and IFN-γ for intact HIV DNA (**A**) and IFN-β and IL-10 for defective HIV DNA (**B**), based on associations identified in Models 1 and 2. Values are estimated changes in reservoir decay slope (log2 copies per million CD4+ T cells per week; negative values indicate faster decay) with 95% confidence intervals and p-values. Estimates with p<0.05 are shown in bold.

**A. HIV Intact DNA – first 52 weeks on ART**

| **Cytokine Measurement Timepoint** | **Subsequent  Reservoir Interval** | **IL-10** | **TNF-α** | **IL-17A** | **IL-18** | **IFN-γ** |
| --- | --- | --- | --- | --- | --- | --- |
| Week 0 | Weeks 0-4 | -0.041 (-0.085, 0.003); 0.067 | **-0.11 (-0.2,  -0.02); 0.017** | -0.015 (-0.07, 0.04); 0.59 | -0.071 (-0.16, 0.016); 0.11 | **-0.065 (-0.11,  -0.019); 0.005** |
|  | Weeks 4-24 | 0 (-0.007, 0.007); 0.99 | -0.005 (-0.017, 0.007); 0.42 | -0.007 (-0.015, 0.001); 0.068 | 0 (-0.011, 0.011); 0.99 | 0 (-0.006, 0.005); 0.89 |
| Week 2 | Weeks 2-4 | -0.025 (-0.11, 0.06); 0.57 | -0.074 (-0.26, 0.11); 0.43 | -0.039 (-0.093, 0.015); 0.16 | -0.06 (-0.19, 0.073); 0.38 | 0.031 (-0.041, 0.1); 0.4 |
|  | Weeks 4-24 | 0.001 (-0.006, 0.008); 0.8 | -0.004 (-0.022, 0.013); 0.62 | -0.002 (-0.006, 0.003); 0.49 | 0.003 (-0.009, 0.015); 0.64 | 0.001 (-0.006, 0.007); 0.83 |
| Week 4 | Weeks 4-24 | 0 (-0.007, 0.008); 0.98 | -0.005 (-0.019, 0.009); 0.47 | -0.004 (-0.009, 0); 0.056 | 0 (-0.008, 0.007); 0.97 | 0.002 (-0.003, 0.007); 0.36 |
| Week 8 | Weeks 8-24 | -0.002 (-0.013, 0.009); 0.75 | 0 (-0.018, 0.018); 0.98 | 0.004 (-0.006, 0.014); 0.42 | 0.001 (-0.013, 0.016); 0.85 | 0.001 (-0.009, 0.011); 0.85 |
| Week 12 | Weeks 12-24 | -0.001 (-0.012, 0.01); 0.84 | 0.005 (-0.016, 0.026); 0.67 | 0.003 (-0.004, 0.01); 0.44 | 0.006 (-0.005, 0.017); 0.28 | 0.005 (-0.003, 0.013); 0.24 |
| Week 16 | Weeks 16-24 | -0.003 (-0.016, 0.01); 0.64 | -0.013 (-0.037, 0.011); 0.28 | 0 (-0.012, 0.011); 0.96 | 0.007 (-0.009, 0.022); 0.39 | 0.003 (-0.01, 0.016); 0.64 |
| Week 20 | Weeks 20-24 | 0.003 (-0.009, 0.015); 0.62 | 0.019 (-0.001, 0.039); 0.063 | 0.003 (-0.009, 0.015); 0.6 | 0.007 (-0.012, 0.025); 0.48 | 0.005 (-0.003, 0.013); 0.24 |

**B. HIV Defective DNA – first 52 weeks on ART**

| **Cytokine Measurement Timepoint** | **Subsequent**  **Reservoir Interval** | **IFN-β** | **IL-10** |
| --- | --- | --- | --- |
| Week 0 | Weeks 0-4 | **-0.04 (-0.068, -0.012); 0.006** | 0.018 (-0.056, 0.092); 0.63 |
|  | Weeks 4-24 | 0.004 (-0.001, 0.008); 0.095 | -0.001 (-0.014, 0.013); 0.9 |
| Week 2 | Weeks 2-4 | -0.007 (-0.069, 0.056); 0.83 | 0.094 (-0.042, 0.23); 0.18 |
|  | Weeks 4-24 | 0.001 (-0.005, 0.007); 0.71 | -0.004 (-0.016, 0.009); 0.56 |
| Week 4 | Weeks 4-24 | 0 (-0.004, 0.005); 0.83 | **-0.02 (-0.036, -0.004); 0.015** |
| Week 8 | Weeks 8-24 | 0.002 (-0.004, 0.009); 0.54 | 0.006 (-0.012, 0.025); 0.5 |
| Week 12 | Weeks 12-24 | -0.004 (-0.012, 0.004); 0.33 | -0.004 (-0.023, 0.016); 0.72 |
| Week 16 | Weeks 16-24 | -0.004 (-0.011, 0.004); 0.37 | -0.016 (-0.04, 0.008); 0.19 |
| Week 20 | Weeks 20-24 | -0.002 (-0.013, 0.009); 0.71 | 0.001 (-0.024, 0.026); 0.93 |

**Supplementary Table 13. Complementary analyses of associations between cytokine concentrations at specified timepoints and subsequent HIV reservoir changes.** Three complementary models were used to evaluate whether a participant’s cytokine concentration at time t_a_ was associated with subsequent changes in HIV DNA over specified intervals from t_a_ to t_b_. First, a mixed-effect nonlinear tensor spline model (**A-B**) evaluated the association between cytokine concentration at t_a_ and reservoir decay over t_a_ to t_b_, with adjusted models controlling for timing of ART initiation, pre-ART viral load, initial CD4+ T-cell count, and time-varying CD4:CD8 ratio. We then used linear models to quantify reservoir change over the same intervals using (i) net change between the initial and final HIV DNA measurements (**C-D**) and (ii) average change estimated using all HIV DNA measurements within the interval (**E-F**). To increase the number of observations available for the net-change analyses, week 0 was defined as <4 weeks on ART and week 52 as 48–56 weeks on ART. Models were fit separately for each cytokine and time interval. Estimates or associations with p<0.05 are shown in bold.

**A. HIV Intact DNA – Nonlinear Relationship (p-value)**

| **Model** | **Start (t_a_)** | **Stop (t_b_)** | **IL-10^2^** | **TNF-α**^2^ | **IL-17A**^2^ | **IL-18**^2^ | **IFN-γ**^2^ |
| --- | --- | --- | --- | --- | --- | --- | --- |
| Unadjusted | 0 | 4 | **0.0027** | **0.0039** | 0.1277 | **0.0026** | **0.002** |
|  | 0 | 24 | **0.0426** | **0.0011** | **0.0044** | **0.0467** | **0.0029** |
|  | 0 | 52 | **0.027** | **5.00e-04** | **0.0177** | **0.0094** | **3.00e-04** |
|  | 4 | 24 | 0.5773 | 0.3501 | **0** | 0.2007 | 0.7918 |
|  | 4 | 52 | **0.0322** | 0.0636 | **0** | 0.1742 | 0.3102 |
|  | 24 | 52 | 0.1423 | 0.3143 | 0.5677 | 0.285 | 0.7188 |
| Adjusted^1^ | 0 | 4 | 0.0607 | 0.2174 | 0.9279 | 0.1193 | **0.0257** |
|  | 0 | 24 | 0.0576 | **0.0045** | **0.0335** | 0.1076 | 0.0504 |
|  | 0 | 52 | **0.0023** | **7.00e-04** | 0.0844 | 0.3116 | **0.0033** |
|  | 4 | 24 | 0.5652 | 0.8286 | **0.0027** | 0.4008 | 0.2405 |
|  | 4 | 52 | 0.1805 | 0.4262 | **3.00e-04** | 0.1494 | 0.4932 |
|  | 24 | 52 | **0.0437** | **0.0494** | 0.7364 | 0.1844 | 0.6247 |

^1^Adjusted models control for the timing of ART initiation, pre-ART viral load, initial CD4+ T cell count, and time-varying CD4:CD8 ratio.

^2^Cytokine cell values are p-values for nonlinear terms evaluating the association between cytokine concentrations at t_a_ and reservoir decay over t_a_ to t_b_.

**B. HIV Defective DNA – Nonlinear Relationship (p-value)**

| **Model** | **Start (t_a_)** | **Stop (t_b_)** | **IFN-β**^2^ | **IL-10**^2^ |
| --- | --- | --- | --- | --- |
| Unadjusted | 0 | 4 | **0.0029** | **3.00e-04** |
|  | 0 | 24 | 0.5257 | 0.0626 |
|  | 0 | 52 | 0.2184 | **0.0235** |
|  | 4 | 24 | 0.2872 | 0.8172 |
|  | 4 | 52 | 0.4325 | 0.6424 |
|  | 24 | 52 | 0.7338 | 0.128 |
| Adjusted^1^ | 0 | 4 | 0.1286 | **0.0147** |
|  | 0 | 24 | 0.904 | 0.1319 |
|  | 0 | 52 | 0.4165 | 0.4071 |
|  | 4 | 24 | 0.1773 | 0.9931 |
|  | 4 | 52 | 0.4044 | 0.9626 |
|  | 24 | 52 | 0.4253 | 0.246 |

^1^Adjusted models control for the timing of ART initiation, pre-ART viral load, initial CD4+ T cell count, and time-varying CD4:CD8 ratio.

^2^Cytokine cell values are p-values for nonlinear terms evaluating the association between cytokine concentrations at t_a_ and reservoir decay over t_a_ to t_b_.

**C. HIV Intact DNA – Net Reservoir Changes (effect size)**

| **Model** | **Start (t_a_)** | **Stop (t_b_)** | **IL-10**^2^ | **TNF-α**^2^ | **IL-17A**^2^ | **IL-18**^2^ | **IFN-γ**^2^ |
| --- | --- | --- | --- | --- | --- | --- | --- |
| Unadjusted | 0 | 4 | **-0.045 (-0.075,  -0.014); 0.004** | **-0.083 (-0.14,  -0.026); 0.004** | **-0.052 (-0.091,  -0.014); 0.008** | -0.059 (-0.13, 0.013); 0.108 | **-0.039 (-0.063,  -0.015); 0.001** |
|  | 0 | 24 | **-0.016 (-0.028,  -0.004); 0.009** | **-0.03 (-0.05,  -0.01); 0.003** | **-0.025 (-0.039,  -0.01); 0.001** | -0.017 (-0.039, 0.005); 0.134 | **-0.011 (-0.019,  -0.004); 0.004** |
|  | 0 | 52 | -0.004 (-0.021, 0.013); 0.664 | -0.001 (-0.052, 0.05); 0.973 | -0.026 (-0.063, 0.011); 0.166 | -0.026 (-0.055, 0.004); 0.095 | -0.01 (-0.026, 0.006); 0.213 |
|  | 4 | 24 | -0.013 (-0.038, 0.013); 0.324 | -0.007 (-0.052, 0.037); 0.74 | **-0.016 (-0.026,  -0.006); 0.001** | -0.007 (-0.029, 0.016); 0.554 | -0.004 (-0.019, 0.011); 0.585 |
|  | 4 | 52 | -0.018 (-0.044, 0.008); 0.164 | 0.025 (-0.079, 0.13); 0.635 | -0.007 (-0.019, 0.004); 0.192 | -0.01 (-0.048, 0.028); 0.596 | -0.013 (-0.035, 0.009); 0.26 |
|  | 24 | 52 | -0.009 (-0.027, 0.01); 0.373 | 0.013 (-0.025, 0.05); 0.505 | -0.011 (-0.032, 0.01); 0.305 | 0.006 (-0.019, 0.031); 0.627 | -0.003 (-0.016, 0.011); 0.702 |
| Adjusted^1^ | 0 | 4 | -0.021 (-0.049, 0.007); 0.146 | -0.049 (-0.1, 0.005); 0.078 | **-0.037 (-0.069,  -0.006); 0.02** | -0.013 (-0.083, 0.058); 0.724 | -0.018 (-0.041, 0.006); 0.137 |
|  | 0 | 24 | -0.007 (-0.02, 0.006); 0.308 | **-0.018 (-0.035,  -0.001); 0.037** | **-0.015 (-0.026,  -0.004); 0.01** | -0.006 (-0.025, 0.013); 0.542 | -0.005 (-0.013, 0.003); 0.238 |
|  | 0 | 52 | -0.006 (-0.027, 0.015); 0.581 | -0.01 (-0.065, 0.044); 0.712 | **-0.067 (-0.077,  -0.058); 0** | -0.024 (-0.064, 0.016); 0.246 | **-0.01 (-0.015,  -0.006); 0** |
|  | 4 | 24 | 0.003 (-0.013, 0.02); 0.71 | **0.027 (0, 0.054); 0.046** | -0.007 (-0.014, 0.001); 0.069 | 0.004 (-0.011, 0.019); 0.639 | 0.004 (-0.006, 0.013); 0.486 |
|  | 4 | 52 | -0.012 (-0.024, 0.001); 0.073 | 0.04 (-0.047, 0.13); 0.363 | **-0.007 (-0.011,  -0.005); 0** | 0.029 (-0.002, 0.06); 0.065 | 0 (-0.021, 0.021); 0.989 |
|  | 24 | 52 | -0.025 (-0.051, 0.002); 0.068 | -0.029 (-0.16, 0.1); 0.675 | -0.01 (-0.044, 0.023); 0.55 | 0.005 (-0.082, 0.092); 0.907 | -0.01 (-0.039, 0.019); 0.485 |

^1^Adjusted models control for the timing of ART initiation, pre-ART viral load, initial CD4+ T cell count, and time-varying CD4:CD8 ratio.

^2^Cytokine cell values are estimated changes in net log-2 reservoir decay over t_a_ to t_b_ associated with a two-fold higher cytokine concentration at t_a_, with 95% confidence intervals and p-values.

**D. HIV Defective DNA – Net Reservoir Changes**

| **Model** | **Start (t_a_)** | **Stop (t_b_)** | **IFN-β**^2^ | **IL-10**^2^ |
| --- | --- | --- | --- | --- |
| Unadjusted | 0 | 4 | -0.035 (-0.077, 0.007); 0.101 | -0.097 (-0.2, 0.002); 0.055 |
|  | 0 | 24 | -0.001 (-0.009, 0.007); 0.755 | **-0.025 (-0.044, -0.006); 0.009** |
|  | 0 | 52 | -0.002 (-0.008, 0.004); 0.565 | 0 (-0.016, 0.016); 0.983 |
|  | 4 | 24 | 0.006 (-0.005, 0.018); 0.276 | 0.001 (-0.042, 0.043); 0.976 |
|  | 4 | 52 | -0.004 (-0.014, 0.006); 0.436 | **0.037 (0.023, 0.05); 0** |
|  | 24 | 52 | -0.002 (-0.011, 0.008); 0.749 | 0.002 (-0.027, 0.032); 0.867 |
| Adjusted^1^ | 0 | 4 | -0.015 (-0.053, 0.023); 0.429 | -0.03 (-0.13, 0.066); 0.538 |
|  | 0 | 24 | 0 (-0.006, 0.006); 0.931 | -0.01 (-0.032, 0.011); 0.357 |
|  | 0 | 52 | -0.002 (-0.006, 0.002); 0.243 | -0.004 (-0.019, 0.011); 0.596 |
|  | 4 | 24 | 0.009 (-0.004, 0.022); 0.181 | 0.006 (-0.041, 0.054); 0.798 |
|  | 4 | 52 | -0.019 (-0.053, 0.015); 0.276 | **0.041 (0.033, 0.05); 0** |
|  | 24 | 52 | 0.007 (-0.008, 0.023); 0.335 | -0.008 (-0.045, 0.029); 0.678 |

^1^Adjusted models control for the timing of ART initiation, pre-ART viral load, initial CD4+ T cell count, and time-varying CD4:CD8 ratio.

^2^Cytokine cell values are estimated changes in net log-2 reservoir decay over t_a_ to t_b_ associated with a two-fold higher cytokine concentration at t_a_, with 95% confidence intervals and p-values.

**E. HIV Intact DNA – Average Reservoir Changes**

| **Model** | **Start (t_a_)** | **Stop (t_b_)** | **IL-10**^2^ | **TNF-α**^2^ | **IL-17A**^2^ | **IL-18**^2^ | **IFN-γ**^2^ |
| --- | --- | --- | --- | --- | --- | --- | --- |
| Unadjusted | 0 | 4 | **-0.047 (-0.089,  -0.005); 0.028** | **-0.083 (-0.16,  -0.004); 0.04** | -0.045 (-0.1, 0.01); 0.108 | **-0.094 (-0.18,  -0.01); 0.029** | **-0.05 (-0.082,  -0.019); 0.002** |
|  | 0 | 24 | -0.013 (-0.033, 0.006); 0.191 | -0.016 (-0.055, 0.023); 0.41 | -0.009 (-0.035, 0.018); 0.532 | -0.001 (-0.04, 0.037); 0.95 | -0.01 (-0.026, 0.006); 0.208 |
|  | 0 | 52 | -0.008 (-0.028, 0.011); 0.407 | -0.004 (-0.043, 0.034); 0.819 | -0.004 (-0.031, 0.022); 0.739 | 0.006 (-0.032, 0.044); 0.753 | -0.007 (-0.022, 0.009); 0.383 |
|  | 4 | 24 | 0.002 (-0.018, 0.022); 0.863 | -0.022 (-0.056, 0.012); 0.205 | **-0.01 (-0.02, 0); 0.043** | -0.004 (-0.023, 0.014); 0.643 | 0 (-0.013, 0.013); 0.962 |
|  | 4 | 52 | -0.001 (-0.018, 0.016); 0.901 | -0.023 (-0.051, 0.006); 0.119 | **-0.011 (-0.019,  -0.003); 0.006** | 0.001 (-0.015, 0.017); 0.939 | -0.002 (-0.012, 0.009); 0.784 |
|  | 24 | 52 | **-0.015 (-0.03, 0); 0.044** | -0.018 (-0.051, 0.015); 0.276 | -0.014 (-0.031, 0.004); 0.125 | 0.008 (-0.011, 0.026); 0.405 | -0.007 (-0.019, 0.004); 0.214 |
| Adjusted^1^ | 0 | 4 | -0.021 (-0.062, 0.019); 0.299 | -0.046 (-0.13, 0.034); 0.263 | -0.028 (-0.077, 0.021); 0.264 | -0.073 (-0.16, 0.012); 0.092 | -0.031 (-0.063, 0.002); 0.063 |
|  | 0 | 24 | -0.006 (-0.028, 0.016); 0.582 | -0.009 (-0.049, 0.032); 0.672 | -0.001 (-0.028, 0.025); 0.923 | 0.008 (-0.034, 0.051); 0.702 | -0.003 (-0.021, 0.015); 0.73 |
|  | 0 | 52 | -0.002 (-0.024, 0.02); 0.83 | 0 (-0.041, 0.042); 0.99 | 0.001 (-0.026, 0.028); 0.947 | 0.012 (-0.031, 0.055); 0.582 | -0.002 (-0.02, 0.016); 0.849 |
|  | 4 | 24 | 0.005 (-0.013, 0.023); 0.586 | -0.016 (-0.046, 0.015); 0.312 | -0.006 (-0.015, 0.003); 0.182 | -0.007 (-0.024, 0.011); 0.446 | 0.005 (-0.007, 0.016); 0.438 |
|  | 4 | 52 | 0.002 (-0.013, 0.017); 0.797 | -0.015 (-0.041, 0.01); 0.244 | **-0.008 (-0.016,  -0.001); 0.025** | -0.003 (-0.018, 0.011); 0.675 | 0.003 (-0.007, 0.013); 0.571 |
|  | 24 | 52 | **-0.016 (-0.031,  -0.001); 0.033** | -0.04 (-0.084, 0.003); 0.067 | -0.01 (-0.028, 0.008); 0.262 | -0.003 (-0.023, 0.018); 0.803 | -0.007 (-0.021, 0.006); 0.297 |

^1^Adjusted models control for the timing of ART initiation, pre-ART viral load, initial CD4+ T cell count, and time-varying CD4:CD8 ratio.

^2^Cytokine cell values are estimated changes in average log-2 reservoir decay over t_a_ to t_b_ associated with a two-fold higher cytokine concentration at t_a_, with 95% confidence intervals and p-values.

**F. HIV Defective DNA – Average Reservoir Changes**

| **Model** | **Start (t_a_)** | **Stop (t_b_)** | **IFN-β**^2^ | **IL-10**^2^ |
| --- | --- | --- | --- | --- |
| Unadjusted | 0 | 4 | -0.018 (-0.084, 0.047); 0.579 | -0.092 (-0.24, 0.06); 0.236 |
|  | 0 | 24 | -0.01 (-0.025, 0.006); 0.224 | -0.022 (-0.057, 0.012); 0.206 |
|  | 0 | 52 | -0.008 (-0.024, 0.007); 0.29 | -0.015 (-0.05, 0.021); 0.416 |
|  | 4 | 24 | 0.003 (-0.005, 0.01); 0.499 | -0.001 (-0.028, 0.026); 0.922 |
|  | 4 | 52 | 0.002 (-0.005, 0.009); 0.574 | 0.004 (-0.021, 0.029); 0.749 |
|  | 24 | 52 | 0.001 (-0.013, 0.015); 0.911 | -0.032 (-0.072, 0.008); 0.114 |
| Adjusted^1^ | 0 | 4 | 0 (-0.056, 0.056); 0.996 | 0.002 (-0.14, 0.14); 0.972 |
|  | 0 | 24 | -0.005 (-0.02, 0.01); 0.511 | -0.006 (-0.045, 0.032); 0.744 |
|  | 0 | 52 | -0.004 (-0.02, 0.011); 0.576 | -0.002 (-0.041, 0.037); 0.922 |
|  | 4 | 24 | 0.002 (-0.006, 0.01); 0.619 | -0.003 (-0.031, 0.025); 0.838 |
|  | 4 | 52 | 0.002 (-0.005, 0.009); 0.62 | 0.002 (-0.024, 0.028); 0.88 |
|  | 24 | 52 | 0.001 (-0.012, 0.014); 0.858 | -0.019 (-0.056, 0.018); 0.307 |

^1^Adjusted models control for the timing of ART initiation, pre-ART viral load, initial CD4+ T cell count, and time-varying CD4:CD8 ratio.

^2^Cytokine cell values are estimated changes in average log-2 reservoir decay over t_a_ to t_b_ associated with a two-fold higher cytokine concentration at t_a_, with 95% confidence intervals and p-values.

**Supplementary Table 14. Counterfactual estimates of HIV reservoir size at 24 weeks of ART associated with specified two-fold differences in plasma cytokine concentrations at selected timepoints.** Using Model 3, we estimated average treatment effects (ATEs), defined as model-based counterfactual contrasts in predicted HIV reservoir size at 24 weeks associated with a specified two-fold difference in cytokine concentration at each measurement timepoint. Analyses focused on IL-10, TNF-α, IL-17A, IL-18, and IFN-γ for intact HIV DNA (**A**) and IFN-β and IL-10 for defective HIV DNA (**B**), based on associations identified in Models 1 and 2. Values are estimated ATEs (95% CI); p-value. Estimates with p<0.05 are shown in bold.

**A. HIV Intact DNA – Reservoir size at 24 weeks on ART**

| **Cytokine Measurement Timepoint** | **IL-10** | **TNF-α** | **IL-17A** | **IL-18** | **IFN-γ** |
| --- | --- | --- | --- | --- | --- |
| Week 0 | -0.17 (-0.35, 0.003); 0.054 | **-0.63 (-0.98, -0.28); 0** | -0.2 (-0.42, 0.023); 0.08 | -0.26 (-0.59, 0.067); 0.12 | **-0.24 (-0.41, -0.076); 0.004** |
| Week 2 | -0.079 (-0.26, 0.11); 0.4 | -0.17 (-0.56, 0.22); 0.39 | -0.092 (-0.2, 0.02); 0.11 | -0.033 (-0.31, 0.24); 0.81 | 0.12 (-0.029, 0.27); 0.12 |
| Week 4 | -0.048 (-0.24, 0.15); 0.63 | -0.19 (-0.52, 0.14); 0.27 | **-0.11 (-0.22, -0.014); 0.026** | 0.12 (-0.065, 0.31); 0.2 | 0.066 (-0.056, 0.19); 0.29 |
| Week 8 | -0.12 (-0.41, 0.17); 0.42 | -0.14 (-0.63, 0.34); 0.57 | 0.092 (-0.15, 0.33); 0.45 | -0.039 (-0.34, 0.26); 0.8 | 0.079 (-0.13, 0.29); 0.47 |
| Week 12 | -0.017 (-0.19, 0.15); 0.85 | 0.067 (-0.33, 0.46); 0.74 | 0.1 (-0.034, 0.24); 0.14 | 0.037 (-0.21, 0.28); 0.77 | 0.048 (-0.093, 0.19); 0.51 |
| Week 16 | -0.066 (-0.27, 0.13); 0.52 | -0.018 (-0.37, 0.33); 0.92 | -0.035 (-0.19, 0.12); 0.66 | 0.12 (-0.11, 0.34); 0.31 | 0.077 (-0.093, 0.25); 0.37 |
| Week 20 | -0.018 (-0.17, 0.14); 0.82 | 0.043 (-0.19, 0.27); 0.72 | -0.011 (-0.13, 0.11); 0.86 | 0.24 (-0.01, 0.49); 0.06 | 0.034 (-0.061, 0.13); 0.48 |

**B. HIV Defective DNA – Reservoir size at 24 weeks on ART**

| **Cytokine Measurement Timepoint** | **IFN-β** | **IL-10** |
| --- | --- | --- |
| Week 0 | -0.084 (-0.2, 0.033); 0.16 | 0.07 (-0.24, 0.38); 0.66 |
| Week 2 | 0.024 (-0.11, 0.16); 0.73 | 0.092 (-0.21, 0.4); 0.56 |
| Week 4 | 0.086 (-0.031, 0.2); 0.15 | -0.34 (-0.75, 0.074); 0.11 |
| Week 8 | 0.018 (-0.12, 0.16); 0.8 | 0.11 (-0.37, 0.59); 0.66 |
| Week 12 | -0.031 (-0.19, 0.13); 0.7 | -0.038 (-0.33, 0.26); 0.8 |
| Week 16 | **0.12 (0.007, 0.24); 0.037** | -0.32 (-0.68, 0.036); 0.078 |
| Week 20 | 0.046 (-0.094, 0.18); 0.52 | 0.079 (-0.25, 0.41); 0.64 |

**Supplementary Table 15. Counterfactual estimates of HIV reservoir size at 52 weeks of ART associated with specified two-fold differences in plasma cytokine concentrations at selected timepoints.** Using Model 3, we estimated average treatment effects (ATEs), defined as model-based counterfactual contrasts in predicted HIV reservoir size at 52 weeks associated with a specified two-fold difference in cytokine concentration at each measurement timepoint. Analyses focused on IL-10, TNF-α, IL-17A, IL-18, and IFN-γ for intact HIV DNA (**A**) and IFN-β and IL-10 for defective HIV DNA (**B**), based on associations identified in Models 1 and 2. Values are estimated ATEs (95% CI); p-value. Estimates with p<0.05 are shown in bold.

**A. HIV Intact DNA – Reservoir size at 52 weeks on ART**

| **Cytokine Measurement Timepoint** | **IL-10** | **TNF-α** | **IL-17A** | **IL-18** | **IFN-γ** |
| --- | --- | --- | --- | --- | --- |
| Week 0 | -0.17 (-0.46, 0.13); 0.26 | **-0.67 (-1.2, -0.17); 0.009** | **-0.41 (-0.74, -0.076); 0.016** | -0.29 (-0.75, 0.18); 0.22 | **-0.28 (-0.51,  -0.048); 0.018** |
| Week 2 | -0.006 (-0.29, 0.28); 0.97 | -0.36 (-1.1, 0.35); 0.32 | -0.16 (-0.34, 0.027); 0.095 | 0.018 (-0.46, 0.5); 0.94 | 0.097 (-0.16, 0.36); 0.46 |
| Week 4 | 0.004 (-0.36, 0.37); 0.98 | -0.24 (-0.9, 0.42); 0.47 | -0.21 (-0.43, 0.005); 0.055 | -0.007 (-0.37, 0.36); 0.97 | 0.11 (-0.12, 0.35); 0.36 |
| Week 8 | -0.079 (-0.56, 0.4); 0.75 | 0.009 (-0.78, 0.8); 0.98 | 0.18 (-0.25, 0.61); 0.42 | 0.062 (-0.58, 0.7); 0.85 | 0.042 (-0.39, 0.48); 0.85 |
| Week 12 | -0.046 (-0.49, 0.4); 0.84 | 0.18 (-0.65, 1); 0.67 | 0.11 (-0.17, 0.4); 0.44 | 0.25 (-0.19, 0.68); 0.27 | 0.19 (-0.13, 0.51); 0.24 |
| Week 16 | -0.11 (-0.58, 0.36); 0.64 | -0.47 (-1.3, 0.39); 0.28 | -0.01 (-0.43, 0.41); 0.96 | 0.24 (-0.31, 0.79); 0.39 | 0.11 (-0.36, 0.58); 0.64 |
| Week 20 | 0.098 (-0.28, 0.48); 0.62 | 0.6 (-0.029, 1.2); 0.062 | 0.1 (-0.28, 0.49); 0.6 | 0.21 (-0.38, 0.81); 0.48 | 0.16 (-0.11, 0.43); 0.24 |
| Week 24 | **-0.5 (-0.99, -0.018); 0.042** | **-1.2 (-2, -0.26); 0.011** | -0.23 (-0.82, 0.36); 0.45 | -0.38 (-0.93, 0.18); 0.18 | -0.31 (-0.79, 0.18); 0.22 |

**B. HIV Defective DNA – Reservoir size at 52 weeks on ART**

| **Cytokine Measurement Timepoint** | **IFN-β** | **IL-10** |
| --- | --- | --- |
| Week 0 | 0.023 (-0.16, 0.21); 0.8 | 0.031 (-0.52, 0.58); 0.91 |
| Week 2 | 0.042 (-0.2, 0.29); 0.73 | 0.015 (-0.47, 0.5); 0.95 |
| Week 4 | 0.023 (-0.19, 0.24); 0.83 | **-0.95 (-1.7, -0.19); 0.015** |
| Week 8 | 0.091 (-0.2, 0.38); 0.54 | 0.28 (-0.53, 1.1); 0.5 |
| Week 12 | -0.16 (-0.49, 0.16); 0.33 | -0.14 (-0.91, 0.63); 0.72 |
| Week 16 | -0.13 (-0.41, 0.15); 0.37 | -0.58 (-1.4, 0.28); 0.19 |
| Week 20 | -0.068 (-0.43, 0.29); 0.71 | 0.037 (-0.76, 0.83); 0.93 |
| Week 24 | -0.068 (-0.43, 0.29); 0.71 | -0.71 (-1.8, 0.37); 0.2 |

**Supplementary Figure 8. Proposed model illustrating how coordinated immune responses in the first weeks of ART may promote viral control and long-term immune-mediated maintenance of the HIV reservoir.** Sampling timepoints for concurrent plasma cytokine and HIV reservoir measurements are shown on the x-axis (weeks 0, 2, 4, 8, 12, 16, 20, 24, and 52). Our findings highlight the complex, context-dependent relationships of several cytokines – including IL-10, IL-17A, TNF-α, interferons (IFNs), and IL-18 – with reservoir dynamics during acute treated HIV. During the earliest stages of ART, coordinated antiviral, immunoregulatory, and controlled inflammatory responses may contribute to rapid reservoir decay (first two columns). Several of these cytokine pathways also contribute to tissue integrity, suggesting a potential role for mucosal immunity in reservoir establishment and persistence (bottom rows). Together, these findings suggest that balanced immune responses during early ART may favor reservoir decay and contribute to subsequent immune-mediated viral control (third column). In contrast, based on prior studies [34], we hypothesize that during longer-term infection, persistent inflammation, immune exhaustion, and tissue damage may disrupt these pathways despite ART, paralleling patterns observed in autoimmune disease and other chronic infections [60-69] and potentially promoting reservoir persistence and chronic immune dysfunction (fourth column). Green arrows indicate hypothesized favorable immune patterns, whereas red arrows indicate dysregulated patterns. Created with BioRender.com.

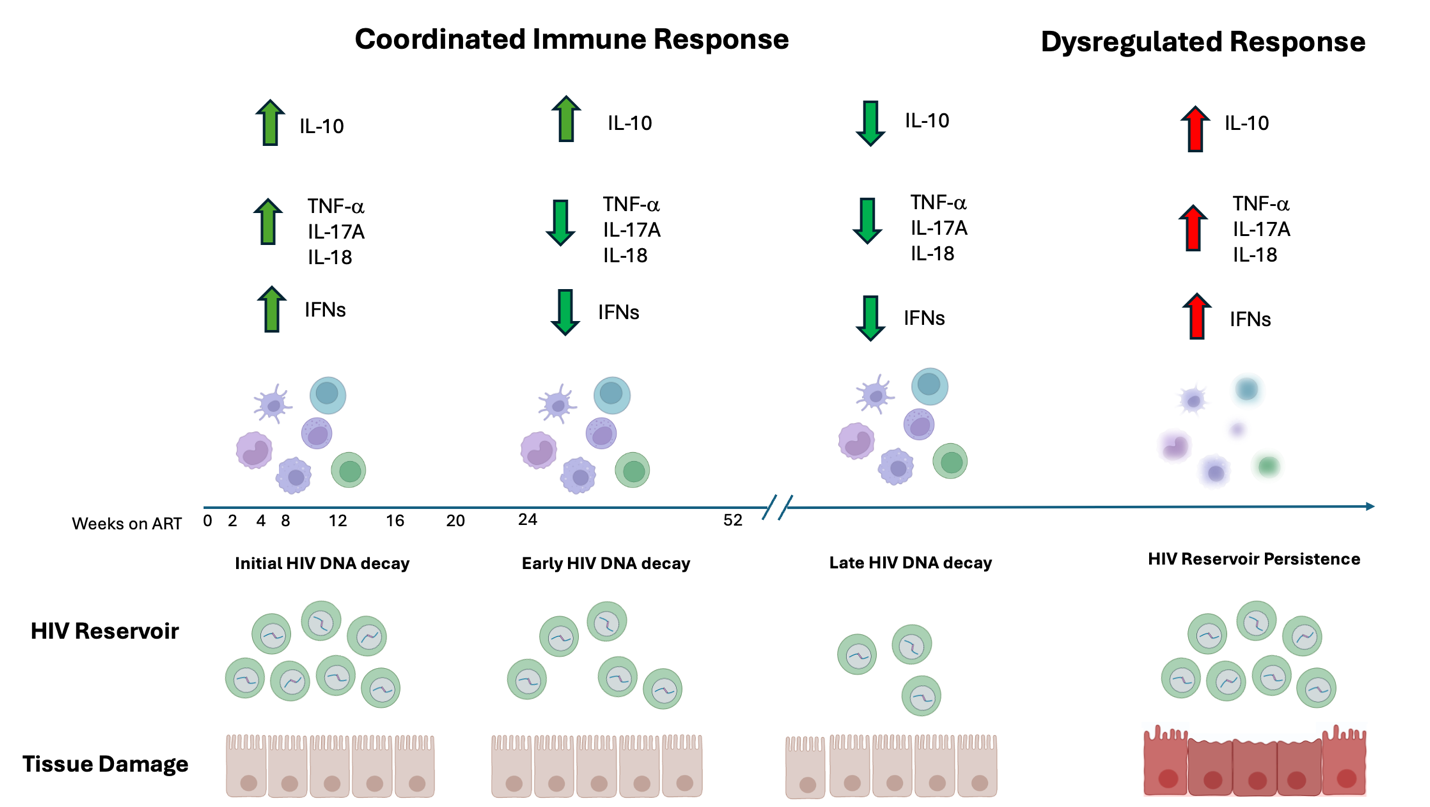
