## Supplementary Methods for "Coordinated Cytokine Responses Are Associated With Rapid HIV Reservoir Decay During Acute Treated HIV Infection"

**METHODS SUPPLEMENT**

**Plasma cytokine quantification and quantifiability analysis**

Plasma concentrations of cytokines (IFN-α2a, IFN-β, IFN-γ, IFN-λ, IL-1β, IL-2, IL-4, IL-6, IL-7, IL-8, IL-9, IL-10, IL-15, IL-17A, IL-18, IL-21, IL-22, IL-27, IL-33, TNF-α, TGF-β1, TGF-β2, and TGF-β3) were quantified at the same timepoints as HIV reservoir DNA (weeks 0, 2, 4, 8, 12, 16, 20, 24, and 52). Using a custom U-Plex multiplexed chemiluminescent assay (Meso Scale Diagnostics), samples were run in duplicate, with concentrations calculated via MSD Discovery Workbench. Samples collected in EDTA or ACD tubes were batch-corrected using ComBat [8]. Values below the assay limit of quantification (BLQ) were reported as zero in the raw MSD data. Eight cytokines were excluded from the primary concentration-based analyses, because >30% of observations were unavailable for quantitative analysis (IFN-α2a, TGF-β3, IL-33, IL-8, IL-2, IL-27, IL-4, and IL-1β). For IL-8, IL-27, and IL-33, the first assay run did not include these analytes; these observations were classified as unmeasured rather than BLQ (**Supplementary Table 2**). As a sensitivity analysis, excluded cytokines were additionally evaluated using a binary quantifiable/BLQ indicator (**Supplementary Table 3**). For the 15 retained cytokines, remaining BLQ values were log-imputed using QRILC [9].

To address whether cytokines excluded from the concentration-based analyses nevertheless showed associations with reservoir decay based on quantifiability, we performed a separate sensitivity analysis. **Model 1** was refit for the excluded cytokines using a binary indicator of quantifiable versus BLQ concentration in place of log2 cytokine concentration. For IL-8, IL-27, and IL-33, observations from the first assay run were treated as unmeasured and excluded rather than classified as BLQ; analyses were therefore restricted to observations in which the analyte had been assayed. IL-27 could not be estimated in this analysis because all available observations were quantifiable when the analyte was assayed. Results are presented in **Supplementary Table 3**.

For the primary concentration-based analyses, cytokine concentrations were standardized within participants, and Spearman correlations of median concentrations by visit were used for hierarchical clustering (Ward’s method). Mixed-effect thin plate splines modeled individual cytokine trajectories relative to baseline, controlling for inter-individual differences, with log-scale fitting and ART initiation constrained to a proportion of one, implemented in R 4.4.1 using mgcv (v1.9-1) with REML estimation [7].

**Cytokine-reservoir change association modeling**

Mixed-effects models were used to evaluate the longitudinal relationship between concurrent plasma cytokine concentrations and HIV reservoir sizes at weeks 0, 2, 4, 8, 12, 16, 20, 24, and 52 weeks. All models were adjusted for initial plasma HIV viral load, baseline CD4 count, and timing of ART initiation, clinical factors that have previously been shown to be associated with HIV reservoir size [1-5]. In addition, a term for the longitudinal CD4:CD8 ratio was included in models to account for potential effects of changes in T-cell composition on the association between cytokines and reservoir decay. Participant-specific random intercepts were included to account for between-individual heterogeneity in reservoir levels. We also evaluated models explicitly adjusting for baseline HIV DNA, including its interaction with time on ART. Cytokine effect estimates were highly concordant with those from models without baseline HIV DNA adjustment (**Supplementary Figure 1;** **Supplementary Tables 5A-B**), indicating that the primary cytokine-reservoir associations were not materially driven by baseline reservoir size. Given this consistency, we selected the more parsimonious specification without baseline HIV DNA adjustment for the final models to limit unnecessary model complexity and overparameterization.

Before incorporating cytokines, we evaluated clinical covariates previously associated with reservoir size [1-5], including pre-ART viral load, baseline CD4+ T cell count, and timing of ART initiation. To account for potential effects of immune cell dynamics on circulating cytokines, we also assessed longitudinal T cell measures (CD4+ count, CD4%, CD8+ count, CD8%, and CD4:CD8 ratio). Among these, the CD4:CD8 ratio demonstrated the strongest association with reservoir decay and was retained in final models (**Supplementary** **Table 4**). Accordingly, all models were adjusted for pre-ART viral load, baseline CD4+ T cell count, timing of ART initiation, and longitudinal CD4:CD8 ratio.

**Models 1 and 2** were used for complementary purposes. We first used the mixed-effect nonlinear model (**Model 2**) to flexibly characterize cytokine-reservoir relationships without imposing a prespecified biphasic linear functional form. We then used the mixed-effect linear spline model (**Model 1**), with the previously identified week-5 inflection point, to quantify and interpret associations within the two established phases of reservoir decay, including changes in decay rate and half-life. Thus, **Model 2** provides a flexible assessment of the overall shape and evidence of cytokine-reservoir associations, whereas **Model 1** provides biologically interpretable phase-specific estimates that are only directly available from the linear spline formulation.

$${log}_{2} y_{it}=\beta_{0}+\beta_{1}D_{i}+\beta_{2}{log}_{2} C_{it}+\beta_{3}R_{it}+\beta_{4}VL_{i}+\beta_{5}CD4_{i}+\left( \beta_{1,0}+\beta_{1,1}D_{i}+\beta_{1,2}{log}_{2} C_{it}+\beta_{1,3}R_{it} \right) \min\left( t,\tau\right)+\left( \beta_{2,0}+\beta_{2,1}D_{i}+\beta_{2,2}{log}_{2} C_{it}+\beta_{2,3}R_{it} \right) \max(0, t-\tau)+\mu_{i}+\epsilon_{it}$$

Model 1

In **Model 1**, subscripts $i$ and $t$ denote unique participants and study visits, respectively. Coefficients on fixed-effect terms are denoted $\beta$. The biphasic linear spline with single knot at $\tau$ is expressed in terms of $\min\left( t,\tau\right)$ and $\max(0, t-\tau)$ for convenient inference within each decay phase. We fixed $\tau=5$ weeks on ART in our models to match the previously published optimal inflection point [6]. The variable $y_{it}$ denotes the HIV reservoir size (copies per million CD4+ T cells), $t$ denotes the number of weeks on ART, $D_{i}$ denotes the delay in ART initiation (weeks), $C_{it}$ denotes the plasma cytokine concentration (pg/mL), $VL_{i}$ denotes the initial log_10_ HIV plasma viral load (copies/mL), $CD4_{i}$ denotes the initial CD4 T cell count (cells/mm^3^), and $R_{\mathrm{it}}$ denotes the CD4:CD8 ratio. The predictors with interaction terms (i.e. $D_{i}$, ${log}_{2} C_{it}$, and $R_{it}$) were centered prior to model fitting. The participant-specific Gaussian random effect is denoted $\mu_{i}$, and the measurement error is denoted $\epsilon_{it}$. Covariates other than $t$ were centered prior to model fitting.

$${log}_{2} y_{it}=ti_{1}\left( t \right)+ti_{2}\left( D_{i} \right)+ti_{3}\left( {log}_{2} C_{it} \right)+ti_{4}\left( R_{it} \right)+ti_{5}\left( t,D_{i} \right)+ti_{6}\left( t,{log}_{2} C_{it} \right)+ti_{7}(t,R_{it})+s_{1}\left( VL_{i} \right)+s_{2}\left( CD4_{i} \right)+\mu_{i}+\epsilon_{it}$$

Model 2

The model terms $ti_{k}$ denote cubic tensor interaction splines and the $s_{k}$ denote cubic regression splines.

A deviance test of nested models was used to assess whether there were significant associations between concurrent cytokine concentrations and changes in HIV reservoir size. In **Model 1**, the null hypothesis tested was $\beta_{2}=\beta_{1,2}=\beta_{2,2}=0$ and in **Model 2**, the null hypothesis tested was ${ti}_{3}\left( \log_{2} C_{it} \right)=0$ and $ti_{6}\left( t,\log_{2} C_{it} \right)=0$; in either instance a significant result (p<0.05) suggested a nontrivial relationship between concurrent plasma cytokine concentrations and longitudinal changes in HIV reservoir size. To further interpret **Model 1** within decay phase $k$, the percent faster decay rate for 2-fold increase in cytokine concentration was estimated as $-100\times\hat{\beta_{k,2}}/\hat{\beta_{k,0}}$ and the half-life was estimated as $\hat{\beta_{k,2}}/\hat{\beta_{k,0}^{2}}$. We note that these measures may not be appropriate if $\hat{\beta_{k,0}}$ is not significantly different from zero. All model fitting was performed with restricted maximum likelihood in R 4.4.1 using the mgcv (v1.9-1) package [7].

**Transformation analysis**

Transformation assumptions were evaluated separately for reservoir outcomes and cytokine predictors. For intact and defective HIV DNA outcomes, mixed-effects Box-Cox analyses retained the participant-specific random intercept and were used to evaluate departures from the log transformation. For cytokine predictors, penalized splines were used to assess whether nonlinear transformations of log2 cytokine concentration improved model fit; results were evaluated after correction for multiple comparisons.

**Multi-cytokine analysis**

All 15 retained log2 cytokines were entered simultaneously into a ridge-penalized version of **Model 1.** In an exploratory extension, all 105 pairwise cytokine interactions were included simultaneously to assess whether the association of one cytokine with reservoir decay varied according to the level of another. Ridge penalties were grouped by cytokine, with interaction terms assigned a combined penalty weight; non-cytokine covariates were not penalized. Penalized terms were standardized to allow comparable shrinkage across terms. Penalties were estimated jointly with the participant-specific random effect using restricted maximum likelihood, allowing the degree of shrinkage to be determined by the data.

Cytokine effect estimates from the multi-cytokine models with and without pairwise interactions were compared using Spearman correlation. Because ridge penalization may attenuate weaker signals, we additionally assessed the sensitivity of the penalized models to effects of different magnitudes by adding a known phase-1 decay-rate effect to the outcome and refitting the model, separately for an individual cytokine and a cytokine pair. This analysis quantified the extent to which imposed effects were recovered after penalization.

**Counterfactual modeling of subsequent reservoir trajectories**

A mixed-effects linear spline model (**Model 3**) was used to examine how cytokine concentrations at specified timepoints related to subsequent reservoir trajectories (**Supplementary Figure 2**). Cytokine terms were restricted to decay phases occurring after the corresponding cytokine measurement, thereby imposing temporal ordering and preventing later cytokine measurement from contributing to earlier reservoir estimates. The model included an interaction term between cytokine concentration and timing of ART initiation to assess effect modification. Model-based contrasts estimated differences in subsequent reservoir size associated with specified two-fold differences in cytokine concentration and were estimated using the multivariate delta method [18]. Predicted reservoir trajectories were computed using the marginaleffects package [19]. Because these data are observational, these estimates were not interpreted as identifying causal effects. All model fitting was performed with restricted maximum likelihood in R 4.4.1 using the mgcv (v1.9-1) package [20].

$${log}_{2} y_{it}=\beta_{0}+\beta_{1}D_{i}+\beta_{2}VL_{i}+\beta_{3}CD4_{i}+\beta_{4}R_{it}+\min\left( t,4 \right)\left( \beta_{1,0}+\beta_{1,1}D_{i}+\beta_{1,4}R_{it} \right)+\max\left( 0,t-4 \right)\left( \beta_{2,0}+{\beta_{2,1}D}_{i}+\beta_{2,4}R_{it}+\beta_{2,5}{log}_{2} C_{i}+{\beta_{2,6}D}_{i}{log}_{2} C_{i} \right)+\mu_{i}+\epsilon_{it}$$

Model 3

The term $C_{i}$ denotes the plasma cytokine concentration (pg/mL) at the specified measurement timepoint (week 4 in this example), and the rest of this model’s terms are defined above; coefficients on fixed-effect terms are denoted β. Cytokine levels in **Model 3** are restricted to interactions following the specified measurement timepoint; thus, the week 4 cytokine concentration cannot contribute to reservoir estimates before week 4, including the intercept. **Model 3** also includes an interaction term between cytokine concentration and timing of ART initiation, $D_{i}C_{i}$, allowing the cytokine-reservoir association to vary with ART timing. The half-life in phase $k$ is $t_{1/2}^{\left( k \right)}(D_{i},R_{it})=-\left( \beta_{k,0}+\beta_{k,1}D_{i}+\beta_{k,4}R_{it} \right)^{-1}$ if the phase is prior to the cytokine measurement and $t_{1/2}^{\left( k \right)}(D_{i},C_{i},R_{it})=-\left( \beta_{2,0}+{\beta_{2,1}D}_{i}+\beta_{2,4}R_{\mathrm{it}}+\beta_{2,5}{log}_{2} C_{i}+{\beta_{2,6}D}_{i}{log}_{2} C_{i} \right)^{-1}$ otherwise; these half-lives are linearly approximated with a Taylor series when necessary. We additionally estimated the average treatment effect (ATE), defined here as the model-based counterfactual contrast in predicted HIV reservoir size at week $t>4$ associated with a specified two-fold difference in cytokine concentration at week 4. For the week 4 model, this contrast is $ATE\left( t \right)=\beta_{2,5}(t-4)$. Estimation and inference of half-lives and ATEs are done with the multivariate delta-method [10]. ATE is retained as the conventional term for this counterfactual estimand. Because cytokine concentrations were observed rather than experimentally assigned, however, estimation of an ATE does not itself establish causal identification. Causal interpretation additionally requires adequate control of confounding and correct model specification, assumptions that cannot be fully verified in this observational cohort. We therefore interpret the estimated ATEs as model-based counterfactual contrasts rather than definitive biological causal effects. All model fitting was performed with restricted maximum likelihood in R 4.4.1 using the mgcv (v1.9-1) package [7]. Predicted HIV reservoir decay curves were computed using the marginaleffects (v0.24.0) package [11]. Prior to model fitting we confirmed substantial inter-individual variability in longitudinal plasma cytokine concentration trends (**Supplementary Table 6**; **Supplementary Figure 3**); this helps to support the predicted decay trajectories for within a reasonable range of hypothetical interventions.

Estimates from **Model 3** were compared with simpler models evaluating whether plasma cytokine concentration at time $l$ was associated with subsequent change in HIV reservoir size over a fixed period $[l,u]$. First, we evaluated this relationship by fitting **Model 2** over $t\in\left[ l,u \right]$ while replacing $C_{it}$ for $C_{il}$. The p-value for the interaction term $ti_{5}\left( t,\log_{2} C_{il} \right)$ was used to evaluate whether there was a nontrivial relationship between the cytokine concentration at $l$ and the rate of reservoir decay over the interval $\left[ l,u \right]$. Next, we individually quantified the change in each participant’s HIV reservoir over $[l,u]$ using the net fold-change, calculated as $\left( u-l \right)^{-1}\log_{2} (y_{iu}/y_{il})$, and the average fold-change, defined as the slope estimate when regressing $\log_{2} (y_{it})$ on $t$ for $t\in[l,u]$ (using an ordinary least squares linear regression model). We then regressed each participant’s net or average reservoir fold-change over $[l,u]$ on their log-scale cytokine concentration at $l$ to estimate the magnitude and direction of the dose-response relationship between plasma cytokine concentrations and subsequent HIV reservoir decay rates.

**REFERENCES**

1.	Archin, N.M., et al., *Immediate antiviral therapy appears to restrict resting CD4+ cell HIV-1 infection without accelerating the decay of latent infection.* Proc Natl Acad Sci U S A, 2012. **109**(24): p. 9523-8.

2.	Buzon, M.J., et al., *Long-term antiretroviral treatment initiated at primary HIV-1 infection affects the size, composition, and decay kinetics of the reservoir of HIV-1-infected CD4 T cells.* J Virol, 2014. **88**(17): p. 10056-65.

3.	Peluso, M.J., et al., *Differential decay of intact and defective proviral DNA in HIV-1-infected individuals on suppressive antiretroviral therapy.* JCI Insight, 2020. **5**(4).

4.	Ananworanich, J., et al., *HIV DNA Set Point is Rapidly Established in Acute HIV Infection and Dramatically Reduced by Early ART.* EBioMedicine, 2016. **11**: p. 68-72.

5.	Crowell, T.A., et al., *Virologic failure is uncommon after treatment initiation during acute HIV infection.* AIDS, 2016. **30**(12): p. 1943-50.

6.	Barbehenn, A., et al., *Rapid biphasic decay of intact and defective HIV DNA reservoir during acute treated HIV disease.* Nat Commun, 2024. **15**(1): p. 9966.

7.	Wood, S.N., *Generalized additive models : an introduction with R*. Second edition. ed. Chapman & Hall/CRC texts in statistical science. 2017, Boca Raton: CRC Press/Taylor & Francis Group. xx, 476 pages.

8.	Johnson, W.E., C. Li, and A. Rabinovic, *Adjusting batch effects in microarray expression data using empirical Bayes methods.* Biostatistics, 2007. **8**(1): p. 118-27.

9.	Wei, R., et al., *Missing Value Imputation Approach for Mass Spectrometry-based Metabolomics Data.* Sci Rep, 2018. **8**(1): p. 663.

10.	Lehmann, E.L. and G. Casella, *Theory of point estimation*. Second edition. ed. Springer texts in statistics. 1998, New York: Springer. 1 online resource (616 pages).

11.	Arel-Bundock, V., N. Greifer, and A. Heiss, *How to Interpret Statistical Models Using marginaleffects for R and Python.* Journal of Statistical Software, 2024. **111**(9).
